# Adherence to the test, trace and isolate system: results from a time series of 21 nationally representative surveys in the UK (the COVID-19 Rapid Survey of Adherence to Interventions and Responses [CORSAIR] study)

**DOI:** 10.1101/2020.09.15.20191957

**Authors:** Louise E Smith, Henry WW Potts, Richard Amlȏt, Nicola T Fear, Susan Michie, G James Rubin

## Abstract

**Objectives:** To investigate rates of adherence to the UK’s test, trace and isolate system over time.

**Design:** Time series of cross-sectional online surveys.

**Setting:** Data were collected between 2 March and 5 August 2020.

**Participants:** 42,127 responses from 31,787 people living in the UK, aged 16 years or over, are presented (21 survey waves, n≈2,000 per wave).

**Main outcome measures:** Identification of the key symptoms of COVID-19 (cough, high temperature / fever, and loss of sense of smell or taste), self-reported adherence to self-isolation if symptomatic, requesting an antigen test if symptomatic, intention to share details of close contacts, self-reported adherence to quarantine if alerted that you had been in contact with a confirmed COVID-19 case.

**Results:** Only 48.9% of participants (95% CI 48.2% to 49.7%) identified key symptoms of COVID-19. Self-reported adherence to test, trace and isolate behaviours was low (self-isolation 18.2%, 95% CI 16.4% to 19.9%; requesting an antigen test 11.9%, 95% CI 10.1% to 13.8%; intention to share details of close contacts 76.1%, 95% CI 75.4% to 76.8%; quarantining 10.9%, 95% CI 7.8% to 13.9%) and largely stable over time. By contrast, intention to adhere to protective measures was much higher. Non-adherence was associated with: men, younger age groups, having a dependent child in the household, lower socioeconomic grade, greater hardship during the pandemic, and working in a key sector.

**Conclusions:** Practical support and financial reimbursement is likely to improve adherence. Targeting messaging and policies to men, younger age groups, and key workers may also be necessary.

**WHAT IS ALREADY KNOWN ON THIS TOPIC:**

- Test, trace and isolate systems are one of the cornerstones of COVID-19 recovery strategy.
- The success of the test, trace and isolation system depends on adherence to isolating if symptomatic, getting a test if symptomatic, passing on details of close contacts if infection is confirmed, and quarantining of contacts.
- Rates of adherence to test, trace and isolate behaviours in the UK need to be systematically investigated.

**WHAT THIS STUDY ADDS:**

- Self-reported adherence to test, trace and isolate behaviours is low; intention to carry out these behaviours is much higher.
- Identification of COVID-19 symptoms is also low.
- Practical support and financial reimbursement are likely to improve adherence to test, trace and isolate behaviours.

## INTRODUCTION

In the absence of a vaccine, governments around the world are relying on test, trace and isolate strategies to prevent the spread of COVID-19.(1) Within the UK, guidance for people who may have COVID-19 has evolved over time, but has focused on the need for people with a persistent new onset cough, fever or loss of their sense of taste or smell to: remain at home for at least seven days from the onset of their symptoms; request an antigen test; and provide the details of their close contacts to a dedicated service if the test result is positive. Isolation and quarantine differ, with isolation being the separation of someone who is ill from others who are not ill and quarantine being the separation and restriction of movement of someone who is not yet ill, but who may have been exposed to a contagious disease.(2) Current guidance requires close contacts of people who have tested positive for COVID-19 to enter quarantine and remain at home for 14 days from the time of their contact.(3)

The ability of the test, trace and isolate system to keep rates of infection under control relies on how well people adhere to it.(4, 5) From the point where an index case develops symptoms to the point where their contacts are allowed to emerge from quarantine, there are multiple stages where adherence might break down.(6) A range of factors may affect adherence. These can be categorised using the Capability, Opportunity, Motivation and Behaviour (COM-B) model.(7) Capability encompasses the psychological and physical capacity to engage in a behaviour. It includes, for example, knowledge as to what the appropriate behaviour is and when to enact it. In the context of test, trace and isolate, knowledge of what the symptoms of COVID-19 are among the UK population has previously been shown to be poor.(8, 9) Insufficient knowledge about the purpose of quarantine has also hindered public health efforts in previous emerging infectious disease outbreaks.(10) Opportunity relates to factors outside the person, for example the presence of financial constraints (9, 11) or cramped accommodation (12) that may make remaining at home difficult to achieve and that, in turn, may be associated with socio-economic status or ethnicity. Motivation describes the psychological processes that energise or inhibit a behaviour and includes the perceived risk associated with a disease outbreak,(13) the belief that you could engage in a behaviour if you wanted to (self-efficacy), and worry about contracting COVID-19 or passing it to others. These motivational components may themselves be influenced by whether information received about a pandemic is viewed as credible (14, 15) and whether the individual considers that they are ’immune’ to COVID-19, particularly if they believe that they have already had it.(16)

During the 2009/10 influenza H1N1 (’swine flu’) pandemic, we analysed a series of 39 surveys commissioned by the English Department of Health to identify the factors associated with adherence to recommended behaviours among members of the public.(17-20) We worked with public health stakeholders to prepare a refined set of questions that could be used in any future pandemic.(21) Since the start of the COVID-19 outbreak, we have worked with the English Department of Health and Social Care to develop and analyse a series of weekly cross-sectional surveys tracking relevant behaviours and their potential predictors in the UK public. In this paper, we report data from 21 of these surveys that tracked adherence to the key components of the test, trace and isolate system over time, and investigate variables associated with capability, opportunity and motivation that may be related to adherence to self-isolation if symptomatic; requesting an antigen test if symptomatic; sharing details of close contacts if symptomatic; and quarantining after being alerted that you have been in close contact with a confirmed COVID-19 case. We also investigated variables associated with correctly identifying the symptoms of COVID-19. Identifying the key factors that increase or decrease adherence can be used to inform policies to improve the functioning of the system and help the UK control the outbreak.

## METHOD

### Design

A national series of cross-sectional surveys were conducted by BMG Research on behalf of the Department of Health and Social Care, England since early in the COVID-19 outbreak (data collection started on 28 January 2020). Surveys were conducted weekly until 1 July (wave 23), after which survey waves were fortnightly. For this paper, we use data from surveys conducted between 2 March 2020 (wave 6) and 5 August 2020 (wave 26). Data were collected over a three-day period (Monday to Wednesday) for each survey wave, except for wave 6 (collected over Monday to Thursday) and wave 12 (collected Tuesday to Wednesday).

### Participants

This study reports on 42,127 responses from 31,787 participants. Participants (n≈2,000 per wave) were recruited from two specialist research panel providers, Respondi (n=50,000) and Savanta (n=31,500). Participants in the first seven waves were recruited from Respondi only; subsequent waves included approximately equal numbers from each panel. Participants were eligible for the study if they were aged 16 years or over and lived in the UK. Participants could be involved in more than one wave. If a respondent completed the survey, they were then unable to participate in the following four waves. Due to an error, some people completed waves more often; 133 people (0.4% of our sample) completed nine waves or more. Quotas were applied based on age and gender (combined) and Government Office Region, and reflected targets based on data from the Office for National Statistics.(22) Participants were reimbursed in points which could be redeemed in cash, gift vouchers or charitable donations (up to 70p).

### Study materials

As the outbreak progressed, relevant questions were added to the survey, while questions judged to be less of a priority were removed. Items were initially derived from a set of questions developed in 2014 in preparation for a future influenza pandemic.(23) These items were refined in 2014 in three rounds of qualitative interviews (n=78) and were assessed for test-retest reliability in two telephone surveys (n=621).(21) Additional items were designed to measure behaviours, attitudes, beliefs and consequences pertinent to the COVID-19 pandemic.

See supplementary materials for full items and response options, with a breakdown of which questions were included in individual survey waves.

#### Outcome measures

Identification of COVID-19 symptoms: single question asking participants to identify the most common symptoms of COVID-19; multiple response options were allowed (up to four initially, up to five from wave 18). We coded participants as identifying symptoms of COVID-19 if they selected cough, high temperature / fever and loss of sense of smell or taste.

Self-isolation: self-reported and intended behaviour. We measured self-reported self-isolation in participants who indicated that they had experienced symptoms of COVID-19 (high temperature / fever, cough, or loss or change of sense of smell or taste) in the last seven days. Participants were asked what, if anything, had caused them to leave home since they developed symptoms. We measured intended self-isolation in participants who had not experienced COVID-19 symptoms in the last week. Participants were asked to imagine they developed COVID-19 the next morning and were asked what would cause them to leave home after developing symptoms, if anything.

Requesting an antigen test: self-reported and intended behaviour. Participants who reported COVID-19 symptoms were asked what actions they had taken since developing symptoms. Response options included “I requested a test to confirm whether I have coronavirus”. Participants not reporting COVID-19 symptoms were asked what actions they would take if they were to develop symptoms.

Sharing details of close contacts: Intended behaviour. Participants who had not experienced COVID-19 symptoms in the last seven days were asked to imagine they had tested positive for COVID-19 and had been “prompted by the NHS contact tracing service”. We asked participants how likely they would be to share details of people they had been in close contact with on a five-point scale from “definitely would” to “definitely would not”. We recoded intention to share details of close contacts into a binary variable (probably or definitely would share details vs not sure, probably or definitely would not). Too few participants in our sample indicated that they had requested a test and tested positive (n=8) to be able to conduct analyses on rates of actual self-reported behaviour.

Quarantining after being alerted: self-reported and intended behaviour. Participants who indicated they had been alerted by a contact tracing service and told they had been in close contact with a confirmed coronavirus case were asked for what reasons, if any, they had left their home in the 14 days after having been contacted the most recent time they were contacted. Participants who had not been alerted were asked what would cause them to leave home after being alerted, if anything.

#### Psychological, health and situational factors

Table 1 lists variables according to the COM-B model.

**Table 1.**
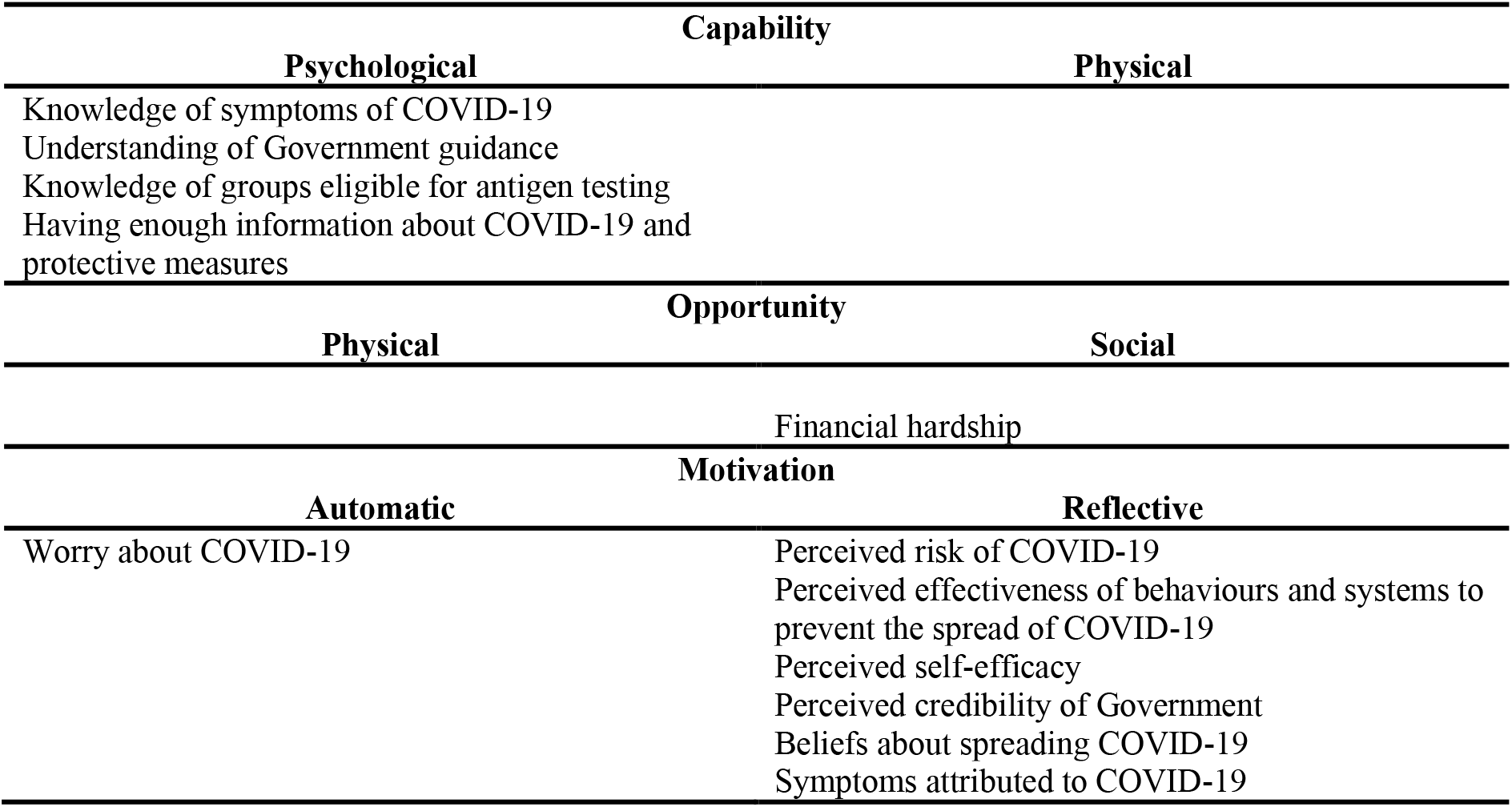
Variables measured classified according to relevance to the COM-B model categories.

Worry and perceived risk: we asked participants how worried they were about COVID-19, and to what extent they thought COVID-19 posed a risk to themselves and others in the UK.

Had COVID-19: participants were asked to state if they thought they “had, or currently have, coronavirus”. Answers were recoded to give a binary variable indicating whether participants thought they had ever had COVID-19.

Symptom attribution: participants who reported having experienced symptoms of COVID-19 in the last week were asked what they thought their symptoms were caused by.

Having enough information about COVID-19 and protective measures: participants were asked to what extent they agreed they had enough information from the Government and other public authorities about a range of topics including symptoms of coronavirus, self-isolation, antigen testing and contact tracing programmes.

Understanding of Government guidance: participants were asked a series of true / false statements about what guidance states you should do if you develop COVID-19 symptoms. Government guidance states that you should not leave home for any reason if you develop symptoms of COVID-19.(3, 24) We coded participants as having incorrect knowledge if they selected any reason for being permitted to leave home.

Knowledge of who was eligible for an antigen test: single item asking participants to select the groups of people eligible for NHS testing if they developed symptoms. Since 18 May 2020, anyone in the UK can have a COVID-19 antigen test if they are symptomatic.(25) We coded participants as having incorrect knowledge if they did not select that everyone who was symptomatic was eligible to be tested.

Perceived credibility of the Government: adapted form of the Meyer Credibility Index (Cronbach’s α=.64).(26)

Perceived effectiveness of behaviours and systems to prevent the spread of COVID-19: we asked participants to what extent they agreed that an effective way to prevent the spread of COVID-19 was to self-isolate, test people with symptoms and to have a contact tracing programme.

Perceived self-efficacy: we asked participants to what extent they agreed that if they wanted to they could self-isolate, book an antigen test online, go to a drive-through testing centre, get a home-testing kit for coronavirus delivered, and return a completed home-testing kit for coronavirus. The period of self-isolation for people with symptoms of COVID-19 was extended from seven days to ten days on 30 July 2020.(27) This change was reflected in data collected on 3 to 5 August (wave 26), but was not analysed here.

Beliefs about spreading COVID-19: we asked participants to what extent they agreed that someone could spread coronavirus to other people even if they did not have symptoms yet, that they were concerned about passing coronavirus on to someone who might be at risk, and that their personal behaviour had an impact on how coronavirus spreads.

Financial hardship: participants were asked to what extent in the past seven days they had been struggling to make ends meet, skipping meals they would usually have, and were finding their current living situation difficult (Cronbach’s α=.74).

#### Personal and clinical characteristics

We asked participants to report their age, gender, employment status, socio-economic grade, highest educational or professional qualification, ethnicity, how many people lived in their household and their marital status. Participants also reported whether: there was a dependent child in the household; they or a household member had a chronic illness; they worked in a key sector; and whether they were self-employed. Participants were asked for their full postcode, from which region and indices of multiple deprivation were determined.(28)

We created a quadratic term for age, to test for a non-linear relationship. We coded participants as having a chronic illness that made them clinically vulnerable to COVID-19 using guidance from the NHS website.(29) Participants were categorised as working in a key sector if they worked in health or social care; education and childcare; key public services; local or national Government; food and essential goods; public safety and national security; transport; or utilities, communication and financial services.(30)

### Ethics

This work was conducted as part of service evaluation of the marketing and communications run by the Department of Health and Social Care, and so did not require ethical approval.

### Patient and public involvement

Lay members served on the advisory group for the project which developed our prototype survey material; this included three rounds of qualitative testing.(21) Due to the rapid nature of this research, the public was not involved in the further development of the survey materials during the COVID-19 pandemic.

### Power

A sample size of 2,000 allows a 95% confidence interval of plus or minus 2% for the prevalence estimate for a survey item with a prevalence of around 50%.

### Analysis

We used logistic regressions to investigate factors associated with: identifying cough, high temperature / fever, and loss of sense of smell or taste (25 May to 5 August 2020); self-isolation (14 April to 5 August 2020); requesting an antigen test (25 May to 5 August 2020); intention to share details of close contacts if tested positive for COVID-19 (1 June to 5 August 2020); and quarantining after being alerted (8 June to 5 August 2020). Although anosmia was added as a symptom of COVID-19 on 18 May 2020,(31) we did not include data collected 18 to 20 May 2020 in these analyses as the announcement happened after data collection had already started.

For each set of analyses, we ran univariable analyses and multivariable analyses. Multivariable regressions investigating factors associated with identification of COVID-19 symptoms, self-isolation and intention to share details of close contacts if tested positive for COVID-19 adjusted for survey wave, region, gender, age (raw and quadratic term), presence of dependent child in the household, being clinically vulnerable to COVID-19, having a household member with a chronic illness, employment status (working vs not working), socio-economic grade (ABC1 vs C2DE), index of multiple deprivation (quartiles), highest educational or professional qualification (degree or higher vs less than degree), ethnicity (coded into six categories), and living alone. Due to small numbers of cases, for analyses investigating factors associated with requesting an antigen test and quarantining after being alerted, it was not advisable to carry out a multivariable regression analysis with a large number of independent variables.(32) Thus, we controlled only for the three factors most strongly associated with the respective outcome in univariable analyses.

Logistic regression analyses were carried out using data from waves starting 14 April to 5 August. 19,441 participants answered just one survey during this period, but 3459 participants (15%) answered more than one survey during this period. Most analyses were conducted on data collected from May or June, therefore the proportion of respondents who answered more than one survey is smaller. Logistic regression analyses treated all responses as independent and did not correct for some participants being in more than one wave. As a sensitivity analysis, we fitted matching generalised estimating equations (GEEs) for some analyses, which adjust for multiple responses. These showed minimal differences in the fitted odds ratios to the logistic regressions.

### Sensitivity analyses

Due to the large number of analyses conducted on each outcome variable (n≈60), we focus our interpretation on those that remained statistically significant after a Bonferroni correction (*p*≤.001).

## RESULTS

As patterns of results were similar for all outcomes, we narratively report factors associated with outcomes at the end of the results section. Full reporting of associations (unadjusted and adjusted) can be found in Tables 2 to 11.

**Table 2.**
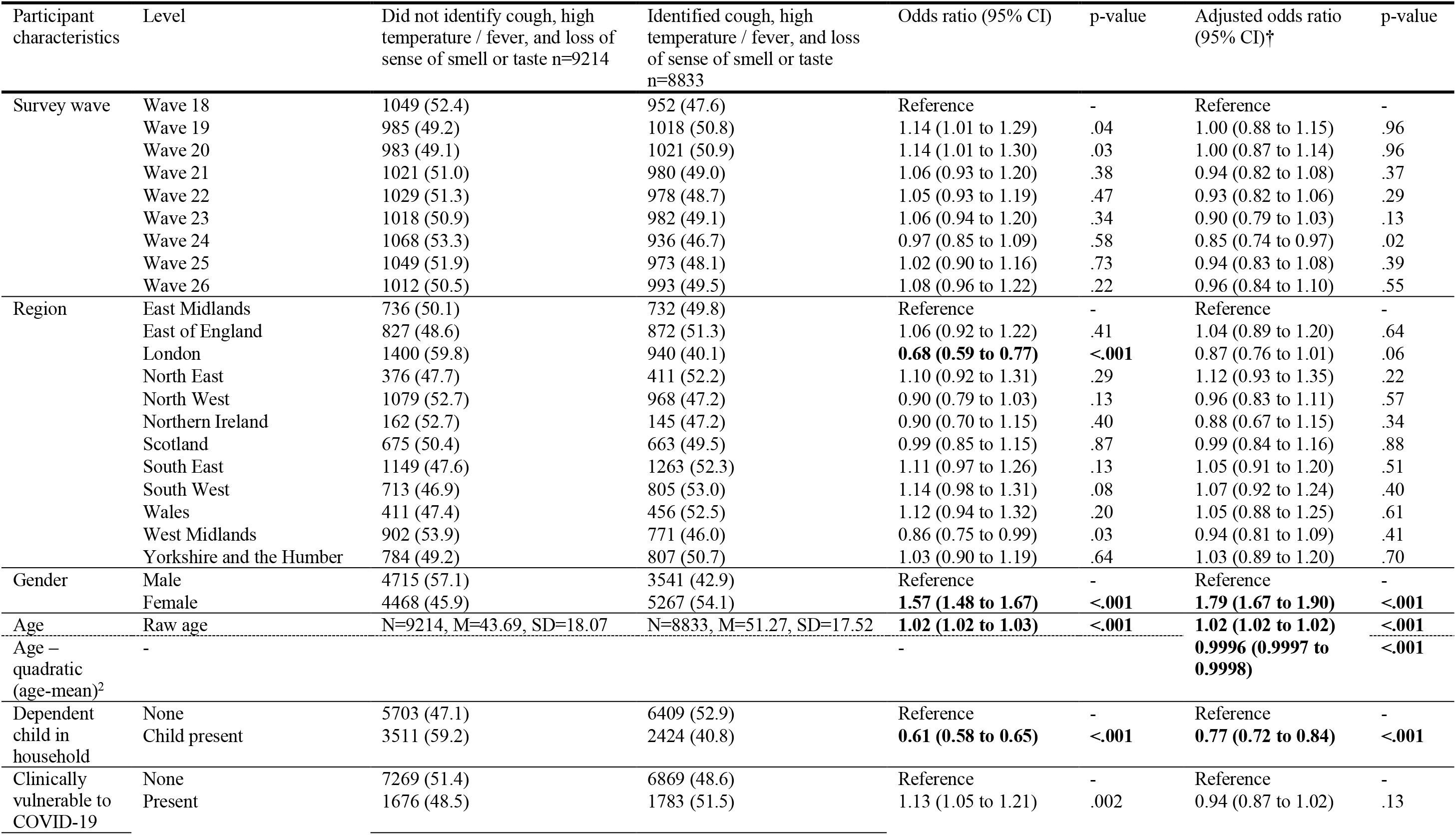
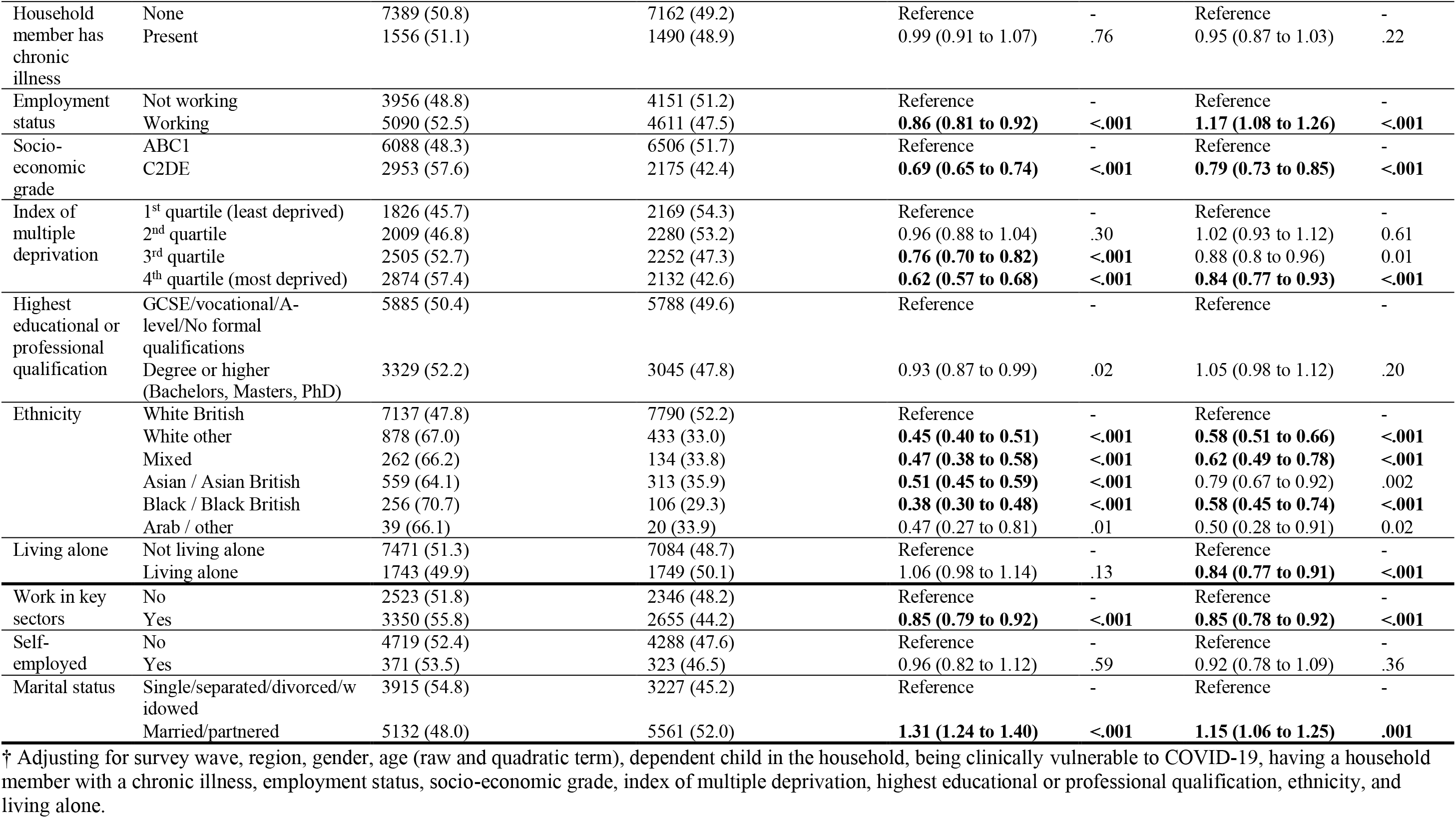
Associations between participant personal and clinical characteristics and correctly identifying cough, high temperature / fever and loss of sense of smell or loss of sense of taste as COVID-19 symptoms. Bolding indicates findings significant at p<0.001.

| Participant characteristics | Level | Did not identify cough, high temperature / fever, and loss of sense of smell or taste n=9214 | Identified cough, high temperature / fever, and loss of sense of smell or taste n=8833 | Odds ratio (95% CI) | p-value | Adjusted odds ratio (95% CI)† | p-value |
| --- | --- | --- | --- | --- | --- | --- | --- |
| Survey wave | Wave 18 | 1049 (52.4) | 952 (47.6) | Reference | - | Reference | - |
|  | Wave 19 | 985 (49.2) | 1018 (50.8) | 1.14 (1.01 to 1.29) | .04 | 1.00 (0.88 to 1.15) | .96 |
|  | Wave 20 | 983 (49.1) | 1021 (50.9) | 1.14 (1.01 to 1.30) | .03 | 1.00 (0.87 to 1.14) | .96 |
|  | Wave 21 | 1021 (51.0) | 980 (49.0) | 1.06 (0.93 to 1.20) | .38 | 0.94 (0.82 to 1.08) | .37 |
|  | Wave 22 | 1029 (51.3) | 978 (48.7) | 1.05 (0.93 to 1.19) | .47 | 0.93 (0.82 to 1.06) | .29 |
|  | Wave 23 | 1018 (50.9) | 982 (49.1) | 1.06 (0.94 to 1.20) | .34 | 0.90 (0.79 to 1.03) | .13 |
|  | Wave 24 | 1068 (53.3) | 936 (46.7) | 0.97 (0.85 to 1.09) | .58 | 0.85 (0.74 to 0.97) | .02 |
|  | Wave 25 | 1049 (51.9) | 973 (48.1) | 1.02 (0.90 to 1.16) | .73 | 0.94 (0.83 to 1.08) | .39 |
|  | Wave 26 | 1012 (50.5) | 993 (49.5) | 1.08 (0.96 to 1.22) | .22 | 0.96 (0.84 to 1.10) | .55 |
| Region | East Midlands | 736 (50.1) | 732 (49.8) | Reference | - | Reference | - |
|  | East of England | 827 (48.6) | 872 (51.3) | 1.06 (0.92 to 1.22) | .41 | 1.04 (0.89 to 1.20) | .64 |
|  | London | 1400 (59.8) | 940 (40.1) | **0.68 (0.59 to 0.77)** | **<.001** | 0.87 (0.76 to 1.01) | .06 |
|  | North East | 376 (47.7) | 411 (52.2) | 1.10 (0.92 to 1.31) | .29 | 1.12 (0.93 to 1.35) | .22 |
|  | North West | 1079 (52.7) | 968 (47.2) | 0.90 (0.79 to 1.03) | .13 | 0.96 (0.83 to 1.11) | .57 |
|  | Northern Ireland | 162 (52.7) | 145 (47.2) | 0.90 (0.70 to 1.15) | .40 | 0.88 (0.67 to 1.15) | .34 |
|  | Scotland | 675 (50.4) | 663 (49.5) | 0.99 (0.85 to 1.15) | .87 | 0.99 (0.84 to 1.16) | .88 |
|  | South East | 1149 (47.6) | 1263 (52.3) | 1.11 (0.97 to 1.26) | .13 | 1.05 (0.91 to 1.20) | .51 |
|  | South West | 713 (46.9) | 805 (53.0) | 1.14 (0.98 to 1.31) | .08 | 1.07 (0.92 to 1.24) | .40 |
|  | Wales | 411 (47.4) | 456 (52.5) | 1.12 (0.94 to 1.32) | .20 | 1.05 (0.88 to 1.25) | .61 |
|  | West Midlands | 902 (53.9) | 771 (46.0) | 0.86 (0.75 to 0.99) | .03 | 0.94 (0.81 to 1.09) | .41 |
|  | Yorkshire and the Humber | 784 (49.2) | 807 (50.7) | 1.03 (0.90 to 1.19) | .64 | 1.03 (0.89 to 1.20) | .70 |
| Gender | Male | 4715 (57.1) | 3541 (42.9) | Reference | - | Reference | - |
|  | Female | 4468 (45.9) | 5267 (54.1) | **1.57 (1.48 to 1.67)** | **<.001** | **1.79 (1.67 to 1.90)** | **<.001** |
| Age | Raw age | N=9214, M=43.69, SD=18.07 | N=8833, M=51.27, SD=17.52 | **1.02 (1.02 to 1.03)** | **<.001** | **1.02 (1.02 to 1.02)** | **<.001** |
| Age – quadratic (age-mean)<sup>2</sup> | - |  |  | - |  | **0.9996 (0.9997 to 0.9998)** | **<.001** |
| Dependent child in household | None | 5703 (47.1) | 6409 (52.9) | Reference | - | Reference | - |
|  | Child present | 3511 (59.2) | 2424 (40.8) | **0.61 (0.58 to 0.65)** | **<.001** | **0.77 (0.72 to 0.84)** | **<.001** |
| Clinically vulnerable to COVID-19 | None | 7269 (51.4) | 6869 (48.6) | Reference | - | Reference | - |
|  | Present | 1676 (48.5) | 1783 (51.5) | 1.13 (1.05 to 1.21) | .002 | 0.94 (0.87 to 1.02) | .13 |
| Household member has chronic illness | None | 7389 (50.8) | 7162 (49.2) | Reference | - | Reference | - |
|  | Present | 1556 (51.1) | 1490 (48.9) | 0.99 (0.91 to 1.07) | .76 | 0.95 (0.87 to 1.03) | .22 |
| Employment status | Not working | 3956 (48.8) | 4151 (51.2) | Reference | - | Reference | - |
|  | Working | 5090 (52.5) | 4611 (47.5) | **0.86 (0.81 to 0.92)** | **<.001** | **1.17 (1.08 to 1.26)** | **<.001** |
| Socio-economic grade | ABC1 | 6088 (48.3) | 6506 (51.7) | Reference | - | Reference | - |
|  | C2DE | 2953 (57.6) | 2175 (42.4) | **0.69 (0.65 to 0.74)** | **<.001** | **0.79 (0.73 to 0.85)** | **<.001** |
| Index of multiple deprivation | 1<sup>st</sup> quartile (least deprived) | 1826 (45.7) | 2169 (54.3) | Reference | - | Reference | - |
|  | 2<sup>nd</sup> quartile | 2009 (46.8) | 2280 (53.2) | 0.96 (0.88 to 1.04) | .30 | 1.02 (0.93 to 1.12) | 0.61 |
|  | 3<sup>rd</sup> quartile | 2505 (52.7) | 2252 (47.3) | **0.76 (0.70 to 0.82)** | **<.001** | 0.88 (0.8 to 0.96) | 0.01 |
|  | 4<sup>th</sup> quartile (most deprived) | 2874 (57.4) | 2132 (42.6) | **0.62 (0.57 to 0.68)** | **<.001** | **0.84 (0.77 to 0.93)** | **<.001** |
| Highest educational or professional qualification | GCSE/vocational/A-level/No formal qualifications | 5885 (50.4) | 5788 (49.6) | Reference | - | Reference | - |
|  | Degree or higher (Bachelors, Masters, PhD) | 3329 (52.2) | 3045 (47.8) | 0.93 (0.87 to 0.99) | .02 | 1.05 (0.98 to 1.12) | .20 |
| Ethnicity | White British | 7137 (47.8) | 7790 (52.2) | Reference | - | Reference | - |
|  | White other | 878 (67.0) | 433 (33.0) | **0.45 (0.40 to 0.51)** | **<.001** | **0.58 (0.51 to 0.66)** | **<.001** |
|  | Mixed | 262 (66.2) | 134 (33.8) | **0.47 (0.38 to 0.58)** | **<.001** | **0.62 (0.49 to 0.78)** | **<.001** |
|  | Asian / Asian British | 559 (64.1) | 313 (35.9) | **0.51 (0.45 to 0.59)** | **<.001** | 0.79 (0.67 to 0.92) | .002 |
|  | Black / Black British | 256 (70.7) | 106 (29.3) | **0.38 (0.30 to 0.48)** | **<.001** | **0.58 (0.45 to 0.74)** | **<.001** |
|  | Arab / other | 39 (66.1) | 20 (33.9) | 0.47 (0.27 to 0.81) | .01 | 0.50 (0.28 to 0.91) | 0.02 |
| Living alone | Not living alone | 7471 (51.3) | 7084 (48.7) | Reference | - | Reference | - |
|  | Living alone | 1743 (49.9) | 1749 (50.1) | 1.06 (0.98 to 1.14) | .13 | **0.84 (0.77 to 0.91)** | **<.001** |
| Work in key sectors | No | 2523 (51.8) | 2346 (48.2) | Reference | - | Reference | - |
|  | Yes | 3350 (55.8) | 2655 (44.2) | **0.85 (0.79 to 0.92)** | **<.001** | **0.85 (0.78 to 0.92)** | **<.001** |
| Self-employed | No | 4719 (52.4) | 4288 (47.6) | Reference | - | Reference | - |
|  | Yes | 371 (53.5) | 323 (46.5) | 0.96 (0.82 to 1.12) | .59 | 0.92 (0.78 to 1.09) | .36 |
| Marital status | Single/separated/divorced/widowed | 3915 (54.8) | 3227 (45.2) | Reference | - | Reference | - |
|  | Married/partnered | 5132 (48.0) | 5561 (52.0) | **1.31 (1.24 to 1.40)** | **<.001** | **1.15 (1.06 to 1.25)** | **.001** |

† Adjusting for survey wave, region, gender, age (raw and quadratic term), dependent child in the household, being clinically vulnerable to COVID-19, having a household member with a chronic illness, employment status, socio-economic grade, index of multiple deprivation, highest educational or professional qualification, ethnicity, and living alone.

**Table 3.**
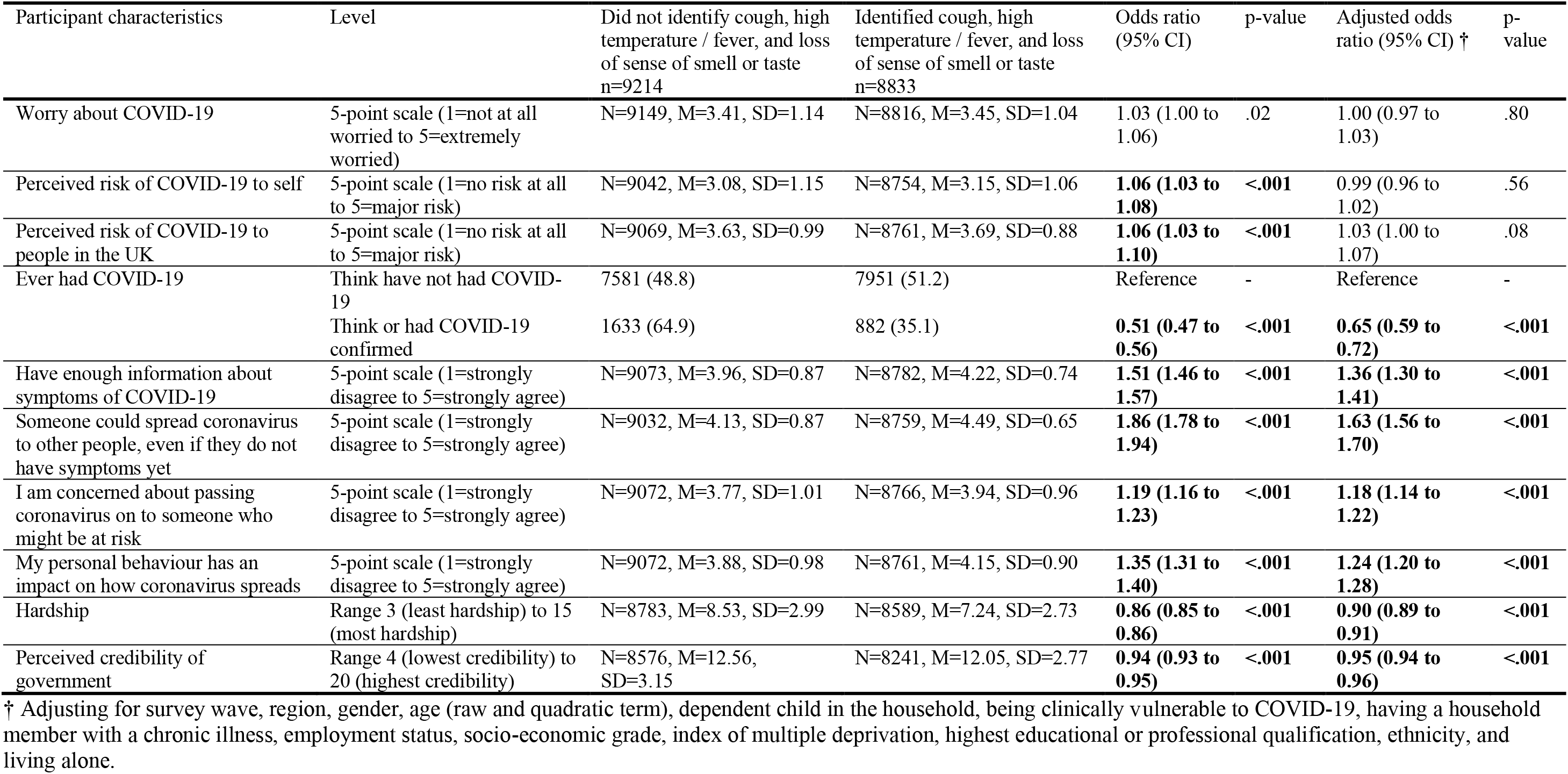
Associations between psychological and situational factors and correctly identifying cough, high temperature / fever and loss of sense of smell or loss of sense of taste as COVID-19 symptoms. Bolding indicates findings significant at p<0.001.

| Participant characteristics | Level | Did not identify cough, high temperature / fever, and loss of sense of smell or taste<br>n=9214 | Identified cough, high temperature / fever, and loss of sense of smell or taste<br>n=8833 | Odds ratio<br>(95% CI) | p-value | Adjusted odds ratio (95% CI) † | p-value |
| --- | --- | --- | --- | --- | --- | --- | --- |
| Worry about COVID-19 | 5-point scale (1=not at all worried to 5=extremely worried) | N=9149, M=3.41, SD=1.14 | N=8816, M=3.45, SD=1.04 | 1.03 (1.00 to 1.06) | .02 | 1.00 (0.97 to 1.03) | .80 |
| Perceived risk of COVID-19 to self | 5-point scale (1=no risk at all to 5=major risk) | N=9042, M=3.08, SD=1.15 | N=8754, M=3.15, SD=1.06 | **1.06 (1.03 to 1.08)** | **<.001** | 0.99 (0.96 to 1.02) | .56 |
| Perceived risk of COVID-19 to people in the UK | 5-point scale (1=no risk at all to 5=major risk) | N=9069, M=3.63, SD=0.99 | N=8761, M=3.69, SD=0.88 | **1.06 (1.03 to 1.10)** | **<.001** | 1.03 (1.00 to 1.07) | .08 |
| Ever had COVID-19 | Think have not had COVID-19 | 7581 (48.8) | 7951 (51.2) | Reference | - | Reference | - |
|  | Think or had COVID-19 confirmed | 1633 (64.9) | 882 (35.1) | **0.51 (0.47 to 0.56)** | **<.001** | **0.65 (0.59 to 0.72)** | **<.001** |
| Have enough information about symptoms of COVID-19 | 5-point scale (1=strongly disagree to 5=strongly agree) | N=9073, M=3.96, SD=0.87 | N=8782, M=4.22, SD=0.74 | **1.51 (1.46 to 1.57)** | **<.001** | **1.36 (1.30 to 1.41)** | **<.001** |
| Someone could spread coronavirus to other people, even if they do not have symptoms yet | 5-point scale (1=strongly disagree to 5=strongly agree) | N=9032, M=4.13, SD=0.87 | N=8759, M=4.49, SD=0.65 | **1.86 (1.78 to 1.94)** | **<.001** | **1.63 (1.56 to 1.70)** | **<.001** |
| I am concerned about passing coronavirus on to someone who might be at risk | 5-point scale (1=strongly disagree to 5=strongly agree) | N=9072, M=3.77, SD=1.01 | N=8766, M=3.94, SD=0.96 | **1.19 (1.16 to 1.23)** | **<.001** | **1.18 (1.14 to 1.22)** | **<.001** |
| My personal behaviour has an impact on how coronavirus spreads | 5-point scale (1=strongly disagree to 5=strongly agree) | N=9072, M=3.88, SD=0.98 | N=8761, M=4.15, SD=0.90 | **1.35 (1.31 to 1.40)** | **<.001** | **1.24 (1.20 to 1.28)** | **<.001** |
| Hardship | Range 3 (least hardship) to 15 (most hardship) | N=8783, M=8.53, SD=2.99 | N=8589, M=7.24, SD=2.73 | **0.86 (0.85 to 0.86)** | **<.001** | **0.90 (0.89 to 0.91)** | **<.001** |
| Perceived credibility of government | Range 4 (lowest credibility) to 20 (highest credibility) | N=8576, M=12.56, SD=3.15 | N=8241, M=12.05, SD=2.77 | **0.94 (0.93 to 0.95)** | **<.001** | **0.95 (0.94 to 0.96)** | **<.001** |

† Adjusting for survey wave, region, gender, age (raw and quadratic term), dependent child in the household, being clinically vulnerable to COVID-19, having a household member with a chronic illness, employment status, socio-economic grade, index of multiple deprivation, highest educational or professional qualification, ethnicity, and living alone.

**Table 4.** Associations between participant personal and clinical characteristics and self-isolating after developing symptoms of COVID-19. Bolding indicates findings significant at p<0.001.

| Participant characteristics | Level | Did not self-isolate<br>n=1587 | Self-isolated n=352 | Odds ratio (95% CI) | p-value | Adjusted odds ratio<br>(95% CI)† | p-value |
| --- | --- | --- | --- | --- | --- | --- | --- |
| Survey wave | Wave 12 | 108 (80.6) | 26 (19.4) | Reference | - | Reference | - |
|  | Wave 13 | 119 (83.8) | 23 (16.2) | 0.80 (0.43 to 1.49) | .49 | 1.03 (0.52 to 2.05) | .94 |
|  | Wave 14 | 89 (80.2) | 22 (19.8) | 1.03 (0.55 to 1.93) | .94 | 1.40 (0.69 to 2.85) | .35 |
|  | Wave 15 | 93 (80.9) | 22 (19.1) | 0.98 (0.52 to 1.85) | .96 | 0.91 (0.45 to 1.86) | .80 |
|  | Wave 16 | 104 (84.6) | 19 (15.4) | 0.76 (0.4 to 1.45) | .41 | 0.90 (0.43 to 1.87) | .77 |
|  | Wave 17 | 84 (73.7) | 30 (26.3) | 1.48 (0.82 to 2.70) | .20 | 1.34 (0.69 to 2.62) | .39 |
|  | Wave 18 | 139 (79.0) | 37 (21.0) | 1.11 (0.63 to 1.94) | .73 | 0.95 (0.5 to 1.81) | .89 |
|  | Wave 19 | 110 (75.9) | 35 (24.1) | 1.32 (0.75 to 2.34) | .34 | 1.41 (0.74 to 2.66) | .29 |
|  | Wave 20 | 109 (79.6) | 28 (20.4) | 1.07 (0.59 to 1.94) | .83 | 0.69 (0.35 to 1.37) | .29 |
|  | Wave 21 | 111 (86.7) | 17 (13.3) | 0.64 (0.33 to 1.24) | .18 | 0.59 (0.28 to 1.25) | .17 |
|  | Wave 22 | 118 (88.1) | 16 (11.9) | 0.56 (0.29 to 1.11) | .10 | 0.45 (0.21 to 0.93) | .03 |
|  | Wave 23 | 96 (81.4) | 22 (18.6) | 0.95 (0.51 to 1.79) | .88 | 0.91 (0.45 to 1.83) | .78 |
|  | Wave 24 | 99 (83.2) | 20 (16.8) | 0.84 (0.44 to 1.60) | .59 | 0.90 (0.44 to 1.85) | .78 |
|  | Wave 25 | 113 (85.6) | 19 (14.4) | 0.70 (0.37 to 1.33) | .28 | 0.66 (0.32 to 1.33) | .24 |
|  | Wave 26 | 95 (85.6) | 16 (14.4) | 0.70 (0.35 to 1.38) | .30 | 0.62 (0.28 to 1.37) | .24 |
| Region | East Midlands | 106 (82.2) | 23 (17.8) | Reference | - | Reference | - |
|  | East of England | 134 (83.8) | 26 (16.3) | 0.89 (0.48 to 1.66) | .72 | 1.01 (0.51 to 1.99) | .98 |
|  | London | 363 (88.8) | 46 (11.2) | 0.58 (0.34 to 1.01) | .05 | 0.81 (0.44 to 1.49) | .49 |
|  | North East | 47 (70.1) | 20 (29.9) | 1.96 (0.98 to 3.91) | .06 | 1.84 (0.84 to 4.04) | .13 |
|  | North West | 209 (85.3) | 36 (14.7) | 0.79 (0.45 to 1.41) | .43 | 0.86 (0.46 to 1.64) | .66 |
|  | Northern Ireland | 16 (94.1) | 1 (5.9) | 0.29 (0.04 to 2.28) | .24 | 0.2 (0.02 to 1.65) | .14 |
|  | Scotland | 93 (76.9) | 28 (23.1) | 1.39 (0.75 to 2.57) | .30 | 1.06 (0.52 to 2.15) | .88 |
|  | South East | 153 (73.2) | 56 (26.8) | 1.69 (0.98 to 2.91) | .06 | 1.52 (0.82 to 2.81) | .19 |
|  | South West | 89 (74.2) | 31 (25.8) | 1.61 (0.87 to 2.95) | .13 | 1.96 (0.99 to 3.85) | .05 |
|  | Wales | 59 (75.6) | 19 (24.4) | 1.48 (0.75 to 2.95) | .26 | 1.71 (0.79 to 3.67) | .17 |
|  | West Midlands | 175 (84.1) | 33 (15.9) | 0.87 (0.48 to 1.56) | .64 | 1.07 (0.56 to 2.06) | .83 |
|  | Yorkshire and the Humber | 143 (81.3) | 33 (18.8) | 1.06 (0.59 to 1.92) | .84 | 0.94 (0.49 to 1.82) | .86 |
| Gender | Male | 942 (84.6) | 172 (15.4) | Reference | - | Reference | - |
|  | Female | 642 (78.3) | 178 (21.7) | **1.52 (1.20 to 1.92)** | **<.001** | **1.75 (1.34 to 2.29)** | **<.001** |
| Age | Raw age | N=1587, M=33.68,<br>SD=13.03 | N=352, M=41.39,<br>SD=17.05 | **1.03 (1.03 to 1.04)** | **<.001** | **1.02 (1.01 to 1.03)** | **<.001** |
| Age – quadratic (age-mean)<sup>2</sup> | - |  |  | - | - | 0.9997 (0.9993 to 1.0002) | .27 |
| Dependent child in household | None | 549 (72.9) | 204 (27.1) | Reference | - | Reference | - |
|  | Child present | 1038 (87.5) | 148 (12.5) | **0.38 (0.30 to 0.49)** | **<.001** | **0.42 (0.32 to 0.57)** | **<.001** |
| Clinically vulnerable to COVID-19 | None | 1091 (82) | 240 (18) | Reference | - | Reference | - |
|  | Present | 465 (82) | 102 (18) | 1.00 (0.77 to 1.29) | .98 | 0.82 (0.61 to 1.11) | .19 |
| Household member has chronic illness | None | 1188 (80.9) | 280 (19.1) | Reference | - | Reference | - |
|  | Present | 368 (85.6) | 62 (14.4) | 0.71 (0.53 to 0.96) | .03 | 0.76 (0.54 to 1.07) | .11 |
| Employment status | Not working | 468 (76.7) | 142 (23.3) | Reference | - | Reference | - |
|  | Working | 1100 (84.4) | 203 (15.6) | **0.61 (0.48 to 0.77)** | **<.001** | 0.74 (0.55 to 1.00) | .05 |
| Socio-economic grade | ABC1 | 748 (76.4) | 231 (23.6) | Reference | - | Reference | - |
|  | C2DE | 821 (87.6) | 116 (12.4) | **0.46 (0.36 to 0.58)** | **<.001** | **0.51 (0.39 to 0.68)** | **<.001** |
| Index of multiple deprivation | 1<sup>st</sup> quartile (least deprived) | 204 (78.8) | 55 (21.2) | Reference | - | Reference | - |
|  | 2<sup>nd</sup> quartile | 302 (81.2) | 70 (18.8) | 0.86 (0.58 to 1.28) | .45 | 0.85 (0.54 to 1.32) | .47 |
|  | 3<sup>rd</sup> quartile | 458 (81.5) | 104 (18.5) | 0.84 (0.58 to 1.21) | .36 | 1.02 (0.67 to 1.54) | .94 |
|  | 4<sup>th</sup> quartile (most deprived) | 623 (83.5) | 123 (16.5) | 0.73 (0.51 to 1.04) | .09 | 0.97 (0.64 to 1.47) | .89 |
| Highest educational or professional qualification | GCSE/vocational/A-level/No formal qualifications | 728 (76.6) | 222 (23.4) | Reference | - | Reference | - |
|  | Degree or higher (Bachelors, Masters, PhD) | 859 (86.9) | 130 (13.1) | **0.50 (0.39 to 0.63)** | **<.001** | **0.63 (0.48 to 0.83)** | **.001** |
| Ethnicity | White British | 1036 (80.2) | 256 (19.8) | Reference | - | Reference | - |
|  | White other | 288 (88.6) | 37 (11.4) | **0.52 (0.36 to 0.75)** | **.001** | 0.76 (0.49 to 1.17) | .21 |
|  | Mixed | 82 (86.3) | 13 (13.7) | 0.64 (0.35 to 1.17) | .15 | 0.86 (0.43 to 1.72) | .67 |
|  | Asian / Asian British | 113 (76.9) | 34 (23.1) | 1.22 (0.81 to 1.83) | .34 | 1.68 (1.05 to 2.69) | .03 |
|  | Black / Black British | 46 (86.8) | 7 (13.2) | 0.62 (0.27 to 1.38) | .24 | 1.07 (0.45 to 2.57) | .87 |
|  | Arab / other | 15 (88.2) | 2 (11.8) | 0.54 (0.12 to 2.37) | .41 | 0.22 (0.04 to 1.17) | .08 |
| Living alone | Not living alone | 1330 (82.4) | 284 (17.6) | Reference | - | Reference | - |
|  | Living alone | 257 (79.1) | 68 (20.9) | 1.24 (0.92 to 1.67) | .16 | 0.86 (0.59 to 1.23) | .40 |
| Work in key sectors | No | 200 (70.9) | 82 (29.1) | Reference | - | Reference | - |
|  | Yes | 1048 (87.5) | 150 (12.5) | **0.35 (0.26 to 0.48)** | **<.001** | **0.46 (0.32 to 0.66)** | **<.001** |
| Self-employed | No | 971 (83.6) | 191 (16.4) | Reference | - | Reference | - |
|  | Yes | 129 (91.5) | 12 (8.5) | 0.47 (0.26 to 0.87) | .02 | 0.73 (0.35 to 1.50) | .39 |
| Marital status | Single/separated/divorced/widowed | 662 (81) | 155 (19) | Reference | - | Reference | - |
|  | Married/partnered | 848 (81.5) | 192 (18.5) | 0.97 (0.76 to 1.22) | .78 | 0.96 (0.70 to 1.30) | .78 |

† Adjusting for survey wave, region, gender, age (raw and quadratic term), dependent child in the household, being clinically vulnerable to COVID-19, having a household member with a chronic illness, employment status, socio-economic grade, index of multiple deprivation, highest educational or professional qualification, ethnicity, and living alone.

**Table 5.** Associations between participant psychological and situational factors and self-isolating after developing symptoms of COVID-19. Bolding indicates findings significant at p<0.001.

| Participant characteristics | Level | Did not self-isolate<br>n=1587 | Self-isolated<br>n=352 | Odds ratio<br>(95% CI) | p-value | Adjusted odds<br>ratio (95% CI)† | p-value |
| --- | --- | --- | --- | --- | --- | --- | --- |
| Worry about COVID-19 | 5-point scale (1=not at all worried to 5=extremely worried) | N=1577, M=3.77, SD=1.19 | N=349, M=3.79, SD=1.11 | 1.01 (0.91 to 1.11) | .88 | 0.97 (0.86 to 1.10) | .67 |
| Perceived risk of COVID-19 to self | 5-point scale (1=no risk at all to 5=major risk) | N=1575, M=3.43, SD=1.18 | N=346, M=3.43, SD=1.16 | 1.00 (0.91 to 1.11) | .99 | 0.95 (0.85 to 1.07) | .44 |
| Perceived risk of COVID-19 to people in the UK | 5-point scale (1=no risk at all to 5=major risk) | N=1574, M=3.73, SD=1.06 | N=348, M=3.92, SD=0.95 | 1.20 (1.07 to 1.35) | .002 | 1.12 (0.98 to 1.28) | .10 |
| Ever had COVID-19 | Think have not had COVID-19<br>Think or had COVID-19 confirmed | 781 (75.8)<br>806 (88.7) | 249 (24.2)<br>103 (11.3) | Reference<br>**0.40 (0.31 to 0.51)** | -<br>**<.001** | Reference<br>**0.46 (0.35 to 0.61)** | -<br>**<.001** |
| Attribute current symptoms to COVID-19‡ | No<br>Yes | 496 (85.4)<br>136 (84.5) | 85 (14.6)<br>25 (15.5) | Reference<br>1.07 (0.66 to 1.74) | -<br>.78 | Reference<br>1.46 (0.82 to 2.60) | -<br>.19 |
| Have enough information about self-isolation§ | 5-point scale (1=strongly disagree to 5=strongly agree) | N=1059, M=3.82, SD=0.94 | N=235, M=4.01, SD=0.89 | 1.26 (1.07 to 1.48) | .01 | 1.12 (0.93 to 1.34) | .23 |
| Knowledge about government guidance if you develop COVID-19 symptoms | Incorrect<br>Correct | 1431 (86.3)<br>156 (55.5) | 227 (13.7)<br>125 (44.5) | Reference<br>**5.05 (3.84 to 6.64)** | -<br>**<.001** | Reference<br>**3.39 (2.47 to 4.64)** | -<br>**<.001** |
| Identified COVID-19 symptoms | No<br>Yes | 1447 (84.6)<br>140 (61.4) | 264 (15.4)<br>88 (38.6) | Reference<br>**3.45 (2.56 to 4.64)** | -<br>**<.001** | Reference<br>**2.50 (1.73 to 3.60)** | -<br>**<.001** |
| An effective way to prevent the spread of coronavirus is to self-isolate for 7 days (not leaving the home at all), if you develop symptoms | 5-point scale (1=strongly disagree to 5=strongly agree) | N=1568, M=3.74, SD=0.99 | N=346, M=4.13, SD=0.92 | **1.57 (1.37 to 1.80)** | **<.001** | 1.28 (1.10 to 1.50) | .002 |
| Confidence that you could self-isolate for 7 days (not leaving the home at all) | 5-point scale (1=strongly disagree to 5=strongly agree) | N=1476, M=3.78, SD=1.02 | N=326, M=4.17, SD=0.89 | **1.56 (1.36 to 1.79)** | **<.001** | 1.28 (1.10 to 1.50) | .002 |
| Someone could spread coronavirus to other people, even if they do not have symptoms yet | 5-point scale (1=strongly disagree to 5=strongly agree) | N=1575, M=3.82, SD=1.02 | N=344, M=4.13, SD=0.87 | **1.43 (1.25 to 1.63)** | **<.001** | 1.18 (1.02 to 1.37) | .03 |
| I am concerned about passing coronavirus on to someone who might be at risk | 5-point scale (1=strongly disagree to 5=strongly agree) | N=1573, M=3.76, SD=1.04 | N=348, M=3.97, SD=0.98 | **1.23 (1.09 to 1.39)** | **.001** | 1.15 (1.00 to 1.32) | .04 |
| My personal behaviour has an impact on how coronavirus spreads | 5-point scale (1=strongly disagree to 5=strongly agree) | N=1575, M=3.79, SD=1.03 | N=347, M=4.02, SD=0.96 | **1.27 (1.12 to 1.44)** | **<.001** | 1.14 (0.99 to 1.31) | .07 |
| Hardship\|\| | Range 3 (least hardship) to 15 (most hardship) | N=1302, M=10.47, SD=2.58 | N=282, M=9.56, SD=2.81 | **0.88 (0.84 to 0.92)** | **<.001** | **0.91 (0.86 to 0.96)** | **.001** |
| Perceived credibility of government | Range 4 (lowest credibility) to 20<br>(highest credibility) | N=1497, M=14,<br>SD=3.41 | N=328, M=12.56,<br>SD=3.40 | **0.89 (0.86 to**<br>**0.92)** | **<.001** | **0.89 (0.85 to**<br>**0.93)** | **<.001** |

† Adjusting for survey wave, region, gender, age (raw and quadratic term), dependent child in the household, being clinically vulnerable to COVID-19, having a household member with a chronic illness, employment status, socio-economic grade, index of multiple deprivation, highest educational or professional qualification, ethnicity, and living alone.
‡ Item added to survey on 15 June 2020.
§ Item added to survey on 18 May 2020.
|| Item added to survey on 20 April 2020.

**Table 6.**
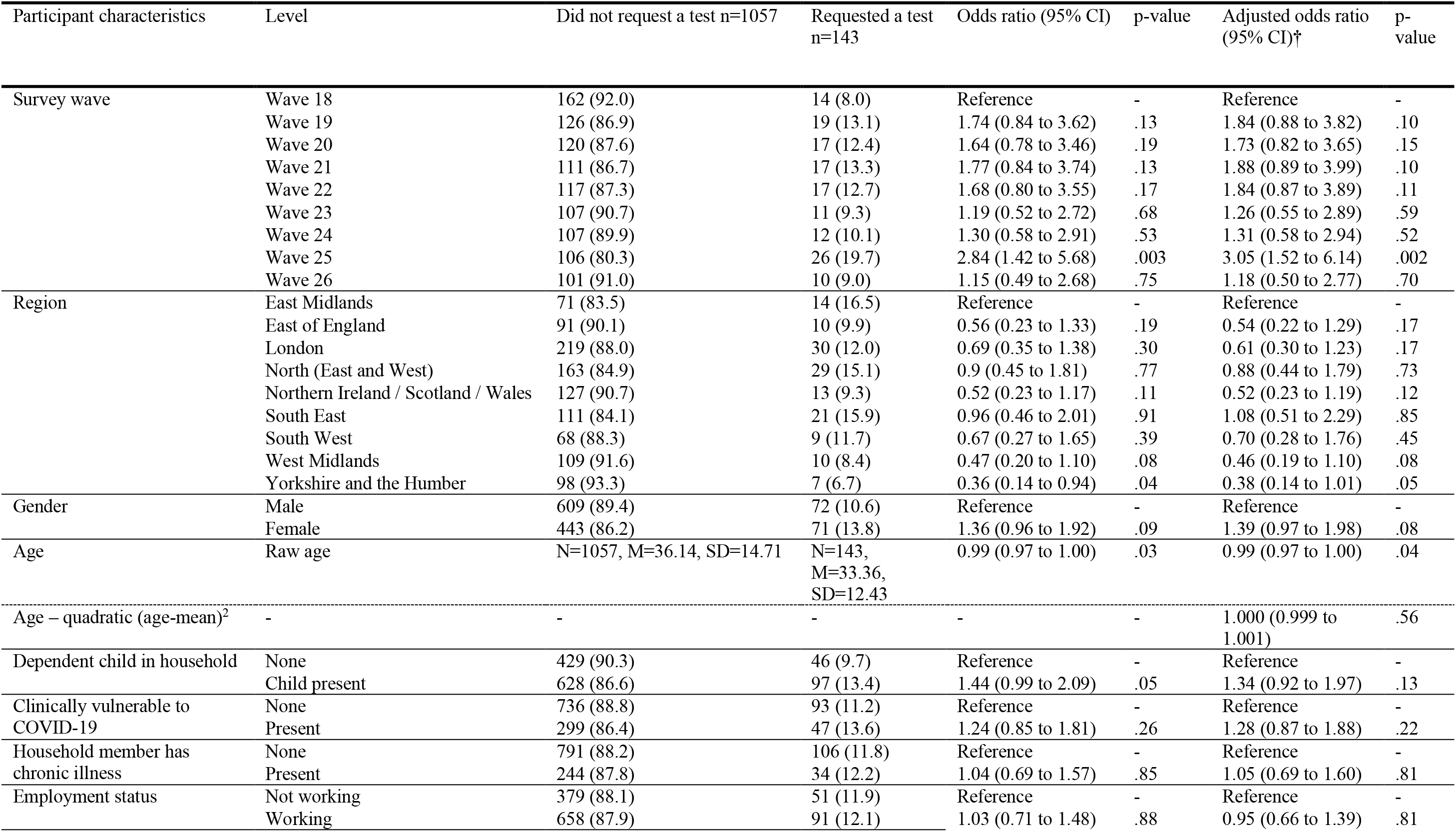
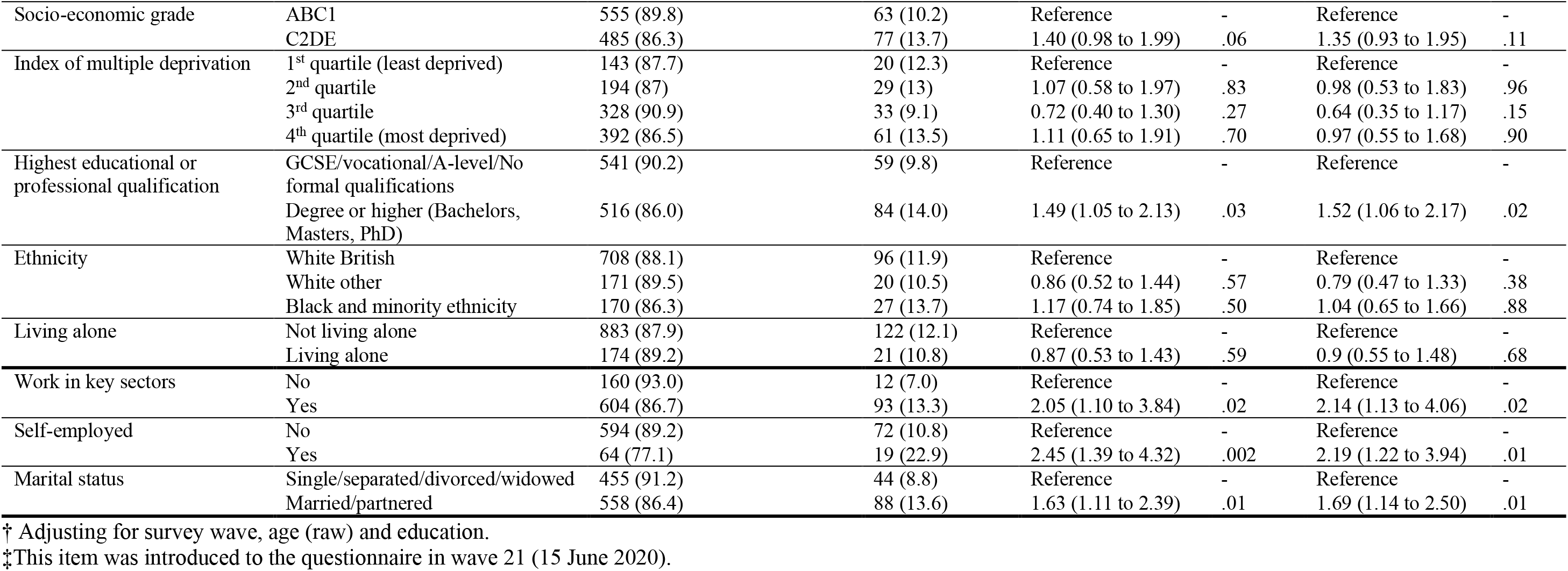
Associations between participant personal and clinical characteristics and requesting an antigen test after developing symptoms of COVID-19. Bolding indicates findings significant at p<0.001.

**Table 7.** Associations between psychological and situational factors and requesting an antigen test after developing symptoms of COVID-19. Bolding indicates findings significant at p<0.001.

| Participant characteristics | Level | Did not request a test n=1057 | Requested a test n=143 | Odds ratio (95% CI) | p-value | Adjusted odds ratio (95% CI)† | p-value |
| --- | --- | --- | --- | --- | --- | --- | --- |
| Worry about COVID-19 | 5-point scale (1=not at all worried to 5=extremely worried) | N=1050, M=3.71, SD=1.19 | N=143, M=3.69, SD=1.05 | 0.98 (0.84 to 1.14) | .78 | 0.98 (0.84 to 1.14) | .80 |
| Perceived risk of COVID-19 to self | 5-point scale (1=no risk at all to 5=major risk) | N=1045, M=3.35, SD=1.17 | N=143, M=3.31, SD=1.19 | 0.97 (0.84 to 1.13) | .71 | 0.98 (0.84 to 1.15) | .84 |
| Perceived risk of COVID-19 to people in the UK | 5-point scale (1=no risk at all to 5=major risk) | N=1047, M=3.70, SD=1.04 | N=142, M=3.56, SD=1.07 | 0.87 (0.74 to 1.03) | .11 | 0.88 (0.74 to 1.04) | .88 |
| Ever had COVID-19 | Think have not had COVID-19 | 610 (88.7) | 78 (11.3) | Reference | - | Reference | - |
|  | Think or had COVID-19 confirmed | 447 (87.3) | 65 (12.7) | 1.14 (0.80 to 1.62) | .47 | 1.09 (0.76 to 1.56) | .65 |
| Attribute current symptoms to COVID-19‡ | No | 511 (88.0) | 70 (12.0) | Reference | - | Reference | - |
|  | Yes | 138 (85.7) | 23 (14.3) | 1.22 (0.73 to 2.02) | .45 | 1.28 (0.77 to 2.15) | .34 |
| Have enough information about testing | 5-point scale (1=strongly disagree to 5=strongly agree) | N=888, M=3.64, SD=1.03 | N=127, M=3.71, SD=1.05 | 1.07 (0.89 to 1.28) | .50 | 1.06 (0.88 to 1.28) | .54 |
| Knowledge about who is eligible to be tested | Incorrect | 710 (88.3) | 94 (11.7) | Reference | - | Reference | - |
|  | Correct | 59 (78.7) | 16 (21.3) | 2.05 (1.13 to 3.71) | .02 | 2.14 (1.17 to 3.92) | .01 |
| Identified COVID-19 symptoms | No | 872 (88.0) | 119 (12.0) | Reference | - | Reference | - |
|  | Yes | 185 (88.5) | 24 (11.5) | 0.95 (0.60 to 1.52) | .83 | 1.13 (0.69 to 1.85) | .63 |
| An effective way to prevent the spread of COVID-19 is to test people with symptoms to confirm whether they have coronavirus | 5-point scale (1=strongly disagree to 5=strongly agree) | N=1039, M=3.84, SD=0.98 | N=141, M=3.81, SD=0.99 | 0.97 (0.81 to 1.15) | .70 | 0.99 (0.83 to 1.19) | .94 |
| Confidence that you could book a test online or via telephone to confirm whether you have coronavirus | 5-point scale (1=strongly disagree to 5=strongly agree) | N=1029, M=3.69, SD=1.02 | N=142, M=3.80, SD=1.06 | 1.11 (0.93 to 1.33) | .23 | 1.11 (0.93 to 1.33) | .25 |
| Confidence that you could go to a drive-through centre to get tested for coronavirus | 5-point scale (1=strongly disagree to 5=strongly agree) | N=1024, M=3.56, SD=1.08 | N=139, M=3.73, SD=1.19 | 1.16 (0.98 to 1.38) | .08 | 1.14 (0.96 to 1.35) | .15 |
| Confidence that you could get a home-testing kit for coronavirus delivered to your home | 5-point scale (1=strongly disagree to 5=strongly agree) | N=1034, M=3.67, SD=1.06 | N=140, M=3.80, SD=1.06 | 1.13 (0.95 to 1.34) | .17 | 1.12 (0.94 to 1.34) | .19 |
| Confidence that you could return a completed home-testing kit for coronavirus via courier (e.g. UPS, Hermes) | 5-point scale (1=strongly disagree to 5=strongly agree) | N=872, M=3.68, SD=1.02 | N=128, M=4.03, SD=0.98 | **1.46 (1.19 to 1.79)** | **<.001** | **1.47 (1.19 to 1.81)** | **<.001** |
| Someone could spread coronavirus to other people, even if they do not have symptoms yet | 5-point scale (1=strongly disagree to 5=strongly agree) | N=1042, M=3.83, SD=0.99 | N=142, M=3.85, SD=1.01 | 1.01 (0.85 to 1.21) | .88 | 1.05 (0.88 to 1.27) | .58 |
| I am concerned about passing coronavirus on to someone who might be at risk | 5-point scale (1=strongly disagree to 5=strongly agree) | N=1043, M=3.75, SD=1.00 | N=143, M=3.78, SD=1.01 | 1.03 (0.86 to 1.23) | .74 | 1.06 (0.89 to 1.26) | .52 |
| My personal behaviour has an impact on how coronavirus spreads | 5-point scale (1=strongly disagree to 5=strongly agree) | N=1049, M=3.78, SD=0.99 | N=142, M=3.77, SD=1.04 | 0.99 (0.83 to 1.18) | .92 | 1.03 (0.86 to 1.23) | .78 |
| Hardship | Range 3 (least hardship) to 15 (most hardship) | N=1005, M=10.36, SD=2.61 | N=138, M=9.88, SD=2.72 | 0.93 (0.87 to 1.00) | .05 | 0.92 (0.86 to 0.99) | .02 |
| Perceived credibility of government | Range 4 (lowest credibility) to 20 (highest credibility) | N=993, M=13.33, SD=3.38 | N=132, M=13.88, SD=3.35 | 1.05 (0.99 to 1.11) | .08 | 1.04 (0.99 to 1.10) | .12 |

† Adjusting for survey wave, age (raw) and education.
‡ This item was introduced to the questionnaire in wave 21 (15 June 2020).

**Table 8.**
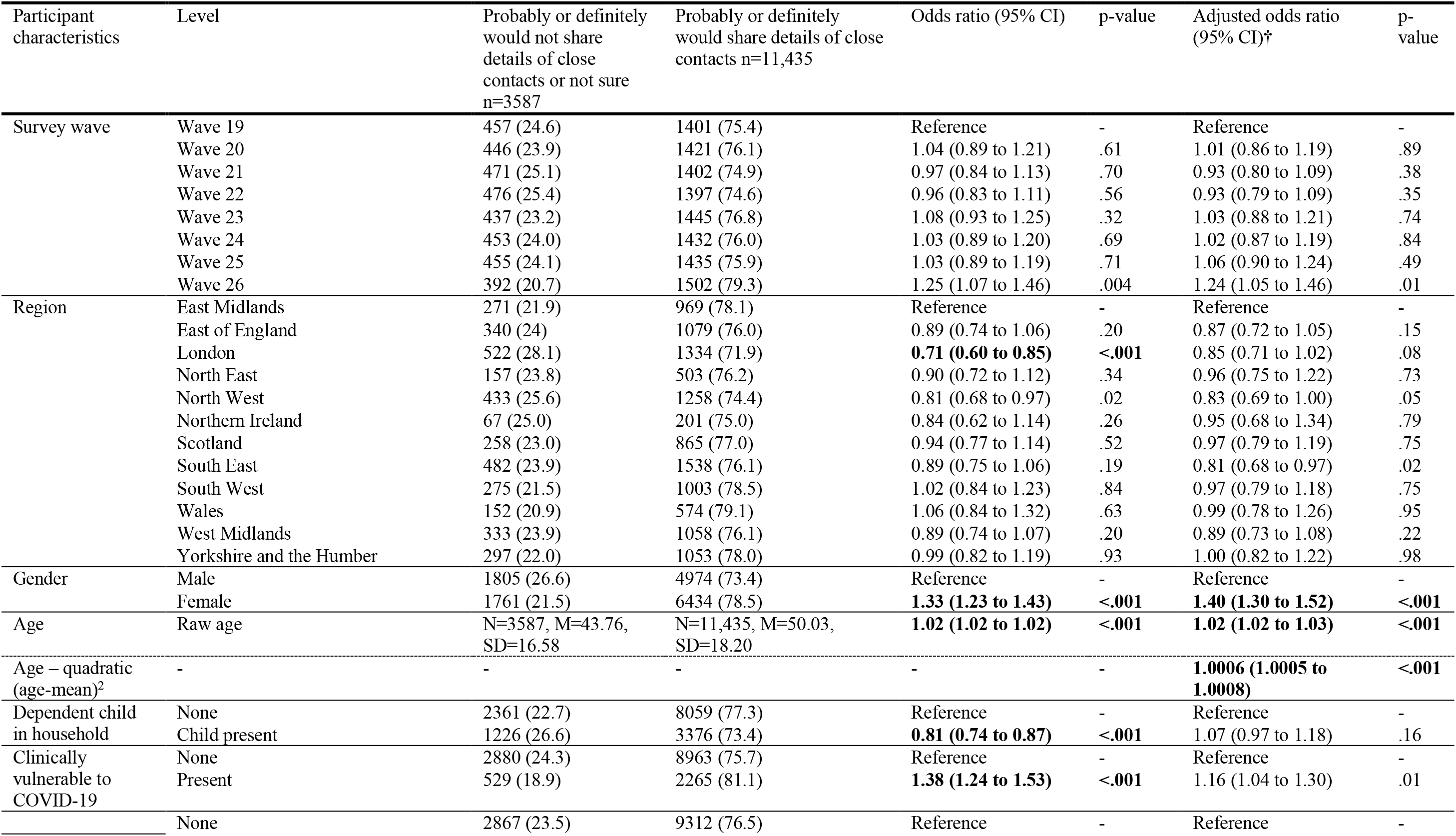
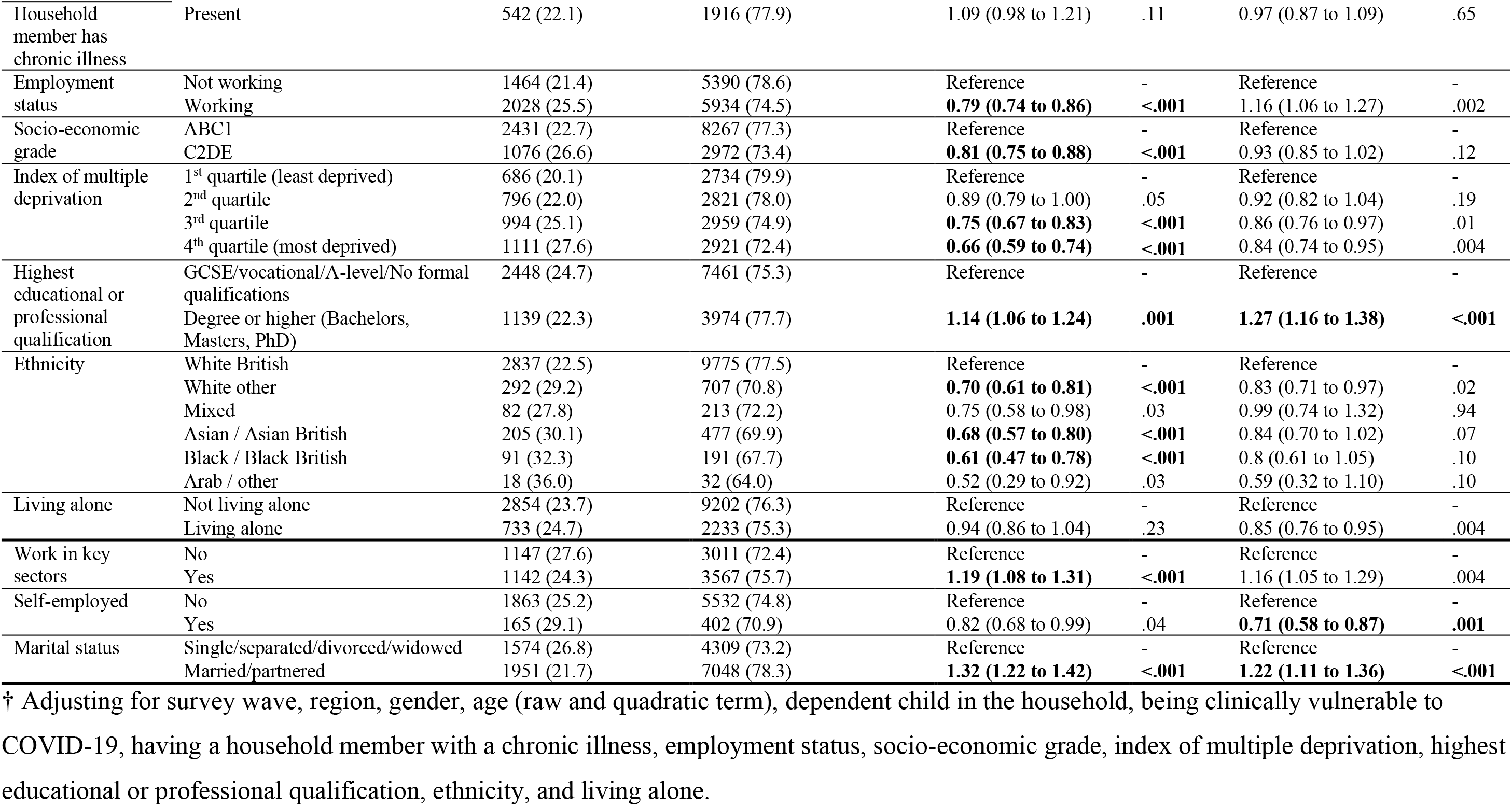
Associations between participant personal and clinical characteristics and intending to share details of your close contacts with the NHS contact tracing service. Bolding indicates findings significant at p<0.001.

**Table 9.** Associations between participant psychological and situational factors and intending to share details of your close contacts with the NHS contact tracing service. Bolding indicates findings significant at p<0.001.

| Participant characteristics | Level | Probably or definitely would not share details of close contacts or not sure<br>n=3587 | Probably or definitely would share details of close contacts<br>n=11,435 | Odds ratio (95% CI) | p-value | Adjusted odds ratio (95% CI)† | p-value |
| --- | --- | --- | --- | --- | --- | --- | --- |
| Worry about COVID-19 | 5-point scale (1=not at all worried to 5=extremely worried) | N=3545, M=3.10, SD=1.19 | N=11,407, M=3.49, SD=1.03 | **1.39 (1.34 to 1.44)** | **<.001** | **1.40 (1.35 to 1.46)** | **<.001** |
| Perceived risk of COVID-19 to self | 5-point scale (1=no risk at all to 5=major risk) | N=3476, M=2.87, SD=1.14 | N=11,334, M=3.16, SD=1.07 | **1.28 (1.24 to 1.33)** | **<.001** | **1.24 (1.20 to 1.29)** | **<.001** |
| Perceived risk of COVID-19 to people in the UK | 5-point scale (1=no risk at all to 5=major risk) | N=3480, M=3.41, SD=1.03 | N=11,357, M=3.72, SD=0.89 | **1.43 (1.37 to 1.49)** | **<.001** | **1.43 (1.37 to 1.49)** | **<.001** |
| Ever had COVID-19 | Think have not had COVID-19 | 3160 (23.8) | 10117 (76.2) | Reference | - | Reference | - |
|  | Think or had COVID-19 confirmed | 427 (24.5) | 1318 (75.5) | 0.96 (0.86 to 1.08) | .54 | 1.15 (1.01 to 1.30) | .03 |
| Have enough information about contact tracing programmes (such as NHS Test and Trace) | 5-point scale (1=strongly disagree to 5=strongly agree) | N=3473, M=3.02, SD=1.11 | N=11,328, M=3.46, SD=1.14 | **1.39 (1.34 to 1.43)** | **<.001** | **1.39 (1.35 to 1.45)** | **<.001** |
| Identified COVID-19 symptoms | No | 2090 (28.6) | 5228 (71.4) | Reference | - | Reference | - |
|  | Yes | 1497 (19.4) | 6207 (80.6) | **1.66 (1.54 to 1.79)** | **<.001** | **1.37 (1.27 to 1.49)** | **<.001** |
| An effective way to prevent the spread of coronavirus is to have a contact tracing programme which anonymously notifies people who have come into close contact with a confirmed coronavirus case to self-isolate | 5-point scale (1=strongly disagree to 5=strongly agree) | N=3353, M=3.11, SD=1.12 | N=11,078, M=4.04, SD=0.93 | **2.31 (2.22 to 2.41)** | **<.001** | **2.29 (2.20 to 2.39)** | **<.001** |
| Someone could spread coronavirus to other people, even if they do not have symptoms yet | 5-point scale (1=strongly disagree to 5=strongly agree) | N=3478, M=4.04, SD=0.89 | N=11,327, M=4.44, SD=0.69 | **1.89 (1.80 to 1.98)** | **<.001** | **1.75 (1.67 to 1.85)** | **<.001** |
| I am concerned about passing coronavirus on to someone who might be at risk | 5-point scale (1=strongly disagree to 5=strongly agree) | N=3489, M=3.56, SD=1.05 | N=11,362, M=3.95, SD=0.95 | **1.48 (1.42 to 1.53)** | **<.001** | **1.52 (1.46 to 1.58)** | **<.001** |
| My personal behaviour has an impact on how coronavirus spreads | 5-point scale (1=strongly disagree to 5=strongly agree) | N=3483, M=3.70, SD=1.00 | N=11,353, M=4.13, SD=0.90 | **1.57 (1.51 to 1.63)** | **<.001** | **1.52 (1.46 to 1.59)** | **<.001** |
| Hardship | Range 3 (least hardship) to 15 (most hardship) | N=3416, M=8.16, SD=2.67 | N=11,060, M=7.54, SD=2.92 | **0.93 (0.92 to 0.94)** | **<.001** | 0.98 (0.96 to 0.99) | .002 |
| Perceived credibility of government | Range 4 (lowest<br>credibility) to 20 (highest<br>credibility) | N=3286,<br>M=11.39,<br>SD=2.95 | N=10,698,<br>M=12.46,<br>SD=2.89 | **1.14 (1.12 to 1.15)** | **<.001** | **1.15 (1.14 to 1.17)** | **<.001** |

† Adjusting for survey wave, region, gender, age (raw and quadratic term), dependent child in the household, being clinically vulnerable to COVID-19, having a household member with a chronic illness, employment status, socio-economic grade, index of multiple deprivation, highest educational or professional qualification, ethnicity, and living alone.

**Table 10.** Associations between participant personal and clinical characteristics and quarantining for 14 days after having been alerted by NHS Test and Trace that you have been in contact with a confirmed COVID-19 case. Bolding indicates findings significant at p<0.001.

| Participant characteristics | Level | Did not quarantine after having been alerted n=361 | Quarantined after having been alerted n=44 | Odds ratio (95% CI) | p-value | Adjusted odds ratio (95% CI)† | p-value |
| --- | --- | --- | --- | --- | --- | --- | --- |
| Survey wave | Wave 20 | 50 (86.2) | 8 (13.8) | Reference | - | Reference | - |
|  | Wave 21 | 52 (91.2) | 5 (8.8) | 0.60 (0.18 to 1.96) | .40 | 0.84 (0.24 to 3.03) | .80 |
|  | Wave 22 | 53 (89.8) | 6 (10.2) | 0.71 (0.23 to 2.18) | .55 | 0.77 (0.22 to 2.61) | .67 |
|  | Wave 23 | 51 (86.4) | 8 (13.6) | 0.98 (0.34 to 2.81) | .97 | 1.09 (0.34 to 3.54) | .88 |
|  | Wave 24 | 51 (89.5) | 6 (10.5) | 0.74 (0.24 to 2.27) | .59 | 0.87 (0.25 to 2.98) | .82 |
|  | Wave 25 | 52 (92.9) | 4 (7.1) | 0.48 (0.14 to 1.70) | .26 | 0.56 (0.14 to 2.18) | .40 |
|  | Wave 26 | 52 (88.1) | 7 (11.9) | 0.84 (0.28 to 2.49) | .76 | 1.10 (0.33 to 3.59) | .88 |
| Region | East Midlands | 27 (93.1) | 2 (6.9) | Reference | - | Reference | - |
|  | East of England | 33 (91.7) | 3 (8.3) | 1.23 (0.19 to 7.88) | .83 | 1.01 (0.14 to 7.11) | .99 |
|  | London | 95 (92.2) | 8 (7.8) | 1.14 (0.23 to 5.67) | .88 | 0.87 (0.16 to 4.60) | .87 |
|  | North (East and West) | 61 (91.0) | 6 (9.0) | 1.33 (0.25 to 7.01) | .74 | 1.32 (0.24 to 7.32) | .75 |
|  | Northern Ireland / Scotland / Wales | 30 (88.2) | 4 (11.8) | 1.80 (0.31 to 10.62) | .52 | 1.41 (0.22 to 9.15) | .72 |
|  | South East | 23 (79.3) | 6 (20.7) | 3.52 (0.65 to 19.17) | .15 | 2.83 (0.46 to 17.34) | .26 |
|  | South West | 17 (81.0) | 4 (19.0) | 3.18 (0.52 to 19.27) | .21 | 3.30 (0.51 to 21.22) | .21 |
|  | West Midlands | 54 (93.1) | 4 (6.9) | 1.00 (0.17 to 5.81) | 1.00 | 1.27 (0.21 to 7.73) | .80 |
|  | Yorkshire and the Humber | 21 (75.0) | 7 (25.0) | 4.5 (0.85 to 23.95) | .08 | 4.33 (0.78 to 24.14) | .10 |
| Gender | Male | 241 (91.3) | 23 (8.7) | Reference | - | Reference | - |
|  | Female | 120 (85.1) | 21 (14.9) | 1.83 (0.98 to 3.45) | .06 | 2.16 (1.08 to 4.31) | .03 |
| Age | Raw age | N=361, M=32.19, SD=10.46 | N=44, M=38.36, SD=16.33 | **1.04 (1.02 to 1.07)** | **.001** | 1.03 (1.01 to 1.05) | .02 |
| Age – quadratic (age-mean)<sup>2</sup> |  | - | - | - | - | 1.000 (0.999 to 1.002) | .52 |
| Dependent child in household | None | 91 (78.4) | 25 (21.6) | Reference | - | Reference | - |
|  | Child present | 270 (93.4) | 19 (6.6) | **0.26 (0.13 to 0.49)** | **<.001** | **0.27 (0.14 to 0.53)** | **<.001** |
| Clinically vulnerable to COVID-19 | None | 239 (86.6) | 37 (13.4) | Reference | - | Reference | - |
|  | Present | 117 (95.1) | 6 (4.9) | 0.33 (0.14 to 0.81) | .02 | 0.32 (0.13 to 0.80) | .02 |
| Household member has chronic illness | None | 299 (89.5) | 35 (10.5) | Reference | - | Reference | - |
|  | Present | 57 (87.7) | 8 (12.3) | 1.20 (0.53 to 2.72) | .66 | 1.18 (0.50 to 2.80) | .71 |
| Employment status | Not working | 103 (87.3) | 15 (12.7) | Reference | - | Reference | - |
|  | Working | 256 (90.1) | 28 (9.9) | 0.75 (0.39 to 1.46) | .40 | 0.80 (0.39 to 1.64) | .55 |
| Socio-economic grade | ABC1 | 173 (88.3) | 23 (11.7) | Reference | - | Reference | - |
|  | C2DE | 188 (90.8) | 19 (9.2) | 0.76 (0.40 to 1.44) | .40 | 0.82 (0.41 to 1.64) | .58 |
| Index of multiple deprivation | 1<sup>st</sup> quartile (least deprived) | 41 (82) | 9 (18) | Reference | - | Reference | - |
|  | 2<sup>nd</sup> quartile | 64 (94.1) | 4 (5.9) | 0.28 (0.08 to 0.99) | .05 | 0.33 (0.09 to 1.25) | .10 |
|  | 3<sup>rd</sup> quartile | 123 (90.4) | 13 (9.6) | 0.48 (0.19 to 1.21) | .12 | 0.51 (0.18 to 1.39) | .19 |
|  | 4<sup>th</sup> quartile (most deprived) | 133 (88.1) | 18 (11.9) | 0.62 (0.26 to 1.48) | .28 | 0.88 (0.33 to 2.34) | .79 |
| Highest educational or professional qualification | GCSE/vocational/A-level/No formal qualifications | 158 (85.4) | 27 (14.6) | Reference | - | Reference | - |
|  | Degree or higher (Bachelors, Masters, PhD) | 203 (92.3) | 17 (7.7) | 0.49 (0.26 to 0.93) | .03 | 0.68 (0.34 to 1.35) | .27 |
| Ethnicity | White British | 221 (86.7) | 34 (13.3) | Reference | - | Reference | - |
|  | White other | 64 (95.5) | 3 (4.5) | 0.30 (0.09 to 1.02) | .05 | 0.34 (0.10 to 1.19) | .09 |
|  | Black and minority ethnicity | 75 (92.6) | 6 (7.4) | 0.52 (0.21 to 1.29) | .16 | 0.39 (0.14 to 1.09) | .07 |
| Living alone | Not living alone | 305 (88.9) | 38 (11.1) | Reference | - | Reference | - |
|  | Living alone | 56 (90.3) | 6 (9.7) | 0.86 (0.35 to 2.13) | .74 | 0.47 (0.17 to 1.28) | .14 |
| Work in key sectors | No | 42 (80.8) | 10 (19.2) | Reference | - | Reference | - |
|  | Yes | 259 (92.8) | 20 (7.2) | 0.32 (0.14 to 0.74) | .01 | 0.41 (0.17 to 0.99) | .05 |
| Self-employed | No | 226 (89.7) | 26 (10.3) | Reference | - | Reference | - |
|  | Yes | 30 (93.8) | 2 (6.3) | 0.58 (0.13 to 2.57) | .47 | 0.72 (0.16 to 3.33) | .67 |
| Marital status | Single/separated/divorced/widowed | 134 (89.3) | 16 (10.7) | Reference | - | Reference | - |
|  | Married/partnered | 214 (88.8) | 27 (11.2) | 1.06 (0.55 to 2.03) | .87 | 1.2 (0.58 to 2.50) | .63 |

† Adjusting for age (raw), dependent child in the household, and being clinically vulnerable to COVID-19.

**Table 11.** Associations between participant psychological and situational factors and quarantining for 14 days after having been alerted by NHS Test and Trace that you have been in contact with a confirmed COVID-19 case. Bolding indicates findings significant at p<0.001.

| Participant characteristics | Level | Did not quarantine after having been alerted n=361 | Quarantined after having been alerted n=44 | Odds ratio (95% CI) | p-value | Adjusted odds ratio (95% CI)<sup>†</sup> | p-value |
| --- | --- | --- | --- | --- | --- | --- | --- |
| Worry about COVID-19 | 5-point scale (1=not at all worried to 5=extremely worried) | N=356, M=3.74, SD=1.17 | N=44, M=3.66, SD=1.08 | 0.95 (0.72 to 1.23) | .68 | 0.97 (0.72 to 1.31) | .84 |
| Perceived risk of COVID-19 to self | 5-point scale (1=no risk at all to 5=major risk) | N=359, M=3.33, SD=1.23 | N=42, M=3.26, SD=0.99 | 0.95 (0.73 to 1.24) | .71 | 0.94 (0.70 to 1.26) | .67 |
| Perceived risk of COVID-19 to people in the UK | 5-point scale (1=no risk at all to 5=major risk) | N=360, M=3.59, SD=1.15 | N=44, M=3.77, SD=0.94 | 1.17 (0.87 to 1.56) | .30 | 1.20 (0.87 to 1.66) | .28 |
| Ever had COVID-19 | Think have not had COVID-19 | 163 (84.9) | 29 (15.1) | Reference | - | Reference | - |
|  | Think or had COVID-19 confirmed | 198 (93.0) | 15 (7.0) | 0.43 (0.22 to 0.82) | .01 | 0.64 (0.32 to 1.31) | .22 |
| Have enough information about contact tracing programmes (such as NHS Test and Trace) | 5-point scale (1=strongly disagree to 5=strongly agree) | N=358, M=3.77, SD=0.94 | N=42, M=3.50, SD=1.06 | 0.76 (0.55 to 1.05) | .09 | 0.85 (0.61 to 1.19) | .34 |
| Identified COVID-19 symptoms | No | 331 (91.4) | 31 (8.6) | Reference | - | Reference | - |
|  | Yes | 30 (69.8) | 13 (30.2) | **4.63 (2.19 to 9.77)** | **<.001** | 2.49 (1.08 to 5.72) | .03 |
| An effective way to prevent the spread of coronavirus is to have a contact tracing programme which anonymously notifies people who have come into close contact with a confirmed coronavirus case to self-isolate | 5-point scale (1=strongly disagree to 5=strongly agree) | N=357, M=3.68, SD=0.96 | N=41, M=3.73, SD=1.18 | 1.05 (0.75 to 1.47) | .77 | 1.21 (0.83 to 1.77) | .32 |
| Confidence that you could self-isolate for 14 days (not leaving the home at all) | 5-point scale (1=strongly disagree to 5=strongly agree) | N=350, M=3.81, SD=0.92 | N=40, M=4.10, SD=0.84 | 1.48 (0.99 to 2.21) | .06 | 1.66 (1.07 to 2.55) | .02 |
| Someone could spread coronavirus to other people, even if they do not have symptoms yet | 5-point scale (1=strongly disagree to 5=strongly agree) | N=357, M=3.77, SD=0.96 | N=43, M=4.07, SD=0.96 | 1.44 (1.00 to 2.09) | .05 | 1.49 (0.99 to 2.23) | .05 |
| I am concerned about passing coronavirus on to someone who might be at risk | 5-point scale (1=strongly disagree to 5=strongly agree) | N=360, M=3.75, SD=0.98 | N=43, M=3.86, SD=0.83 | 1.13 (0.80 to 1.58) | .49 | 1.19 (0.82 to 1.72) | .37 |
| My personal behaviour has an impact on how coronavirus spreads | 5-point scale (1=strongly disagree to 5=strongly agree) | N=359, M=3.73, SD=0.93 | N=43, M=3.74, SD=1.03 | 1.02 (0.72 to 1.43) | .92 | 1.15 (0.80 to 1.65) | .46 |
| Hardship | Range 3 (least hardship) to 15 (most hardship) | N=354, M=10.89, SD=2.52 | N=40, M=9.43, SD=2.63 | **0.81 (0.71 to 0.92)** | **.001** | 0.86 (0.75 to 0.98) | .02 |
| Perceived credibility of government | Range 4 (lowest credibility) to 20 (highest credibility) | N=337, M=14.26, SD=3.13 | N=43, M=12.98, SD=2.52 | 0.87 (0.79 to 0.97) | .01 | 0.90 (0.80 to 1.01) | .09 |

<sup>†</sup> Adjusting for age (raw), dependent child in the household, and being clinically vulnerable to COVID-19.

### Identification of COVID-19 symptoms

Only 48.9% of participants (95% CI 48.2% to 49.7%) identified cough, high temperature / fever and loss of sense of smell or taste as symptoms of COVID-19. This percentage has remained relatively stable over time. There were initial increases at the start of data collection and when loss of sense of smell or taste was introduced into the guidance, but some decline since April/early May (see Figure 1).

**Figure 1.**
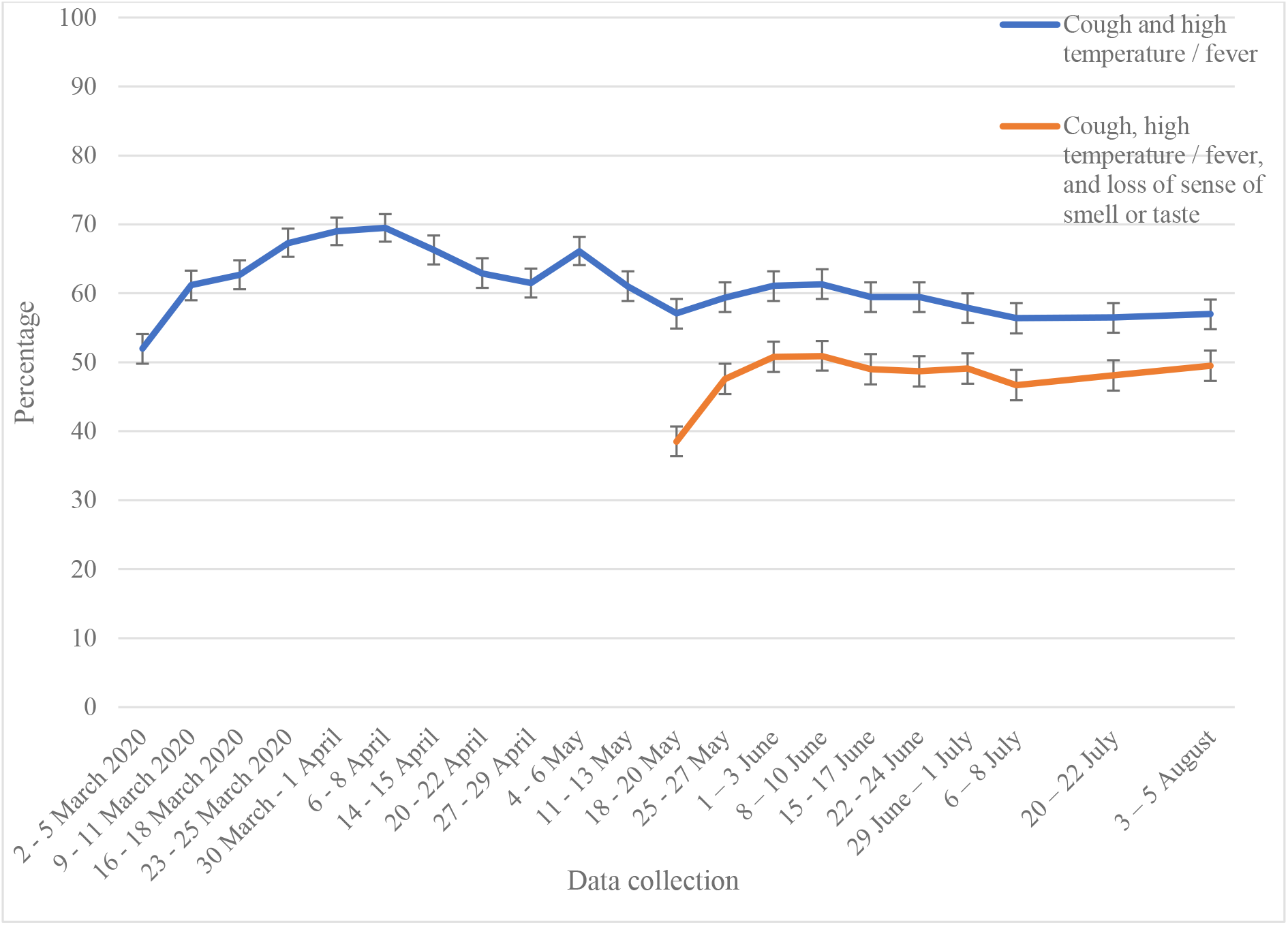
Percentage of people who correctly identified the most common symptoms of COVID-19. Error bars are 95% confidence intervals.

The factors most strongly associated with not identifying COVID-19 symptoms were: male gender; younger age; not identifying as White British; thinking you have had COVID-19; and not knowing that you can spread COVID-19 to others if you are asymptomatic (see Tables 2 and 3).

### Self-isolation

Of those who reported having experienced symptoms of COVID-19 in the last seven days, only 18.2% (95% CI 16.4 to 19.9) said they had not left home since developing symptoms. The percentage of people reporting self-isolating has been largely stable over time (see Figure 2). Intention to self-isolate if you were to develop symptoms of COVID-19 is much higher (around 70%), and has shown a slight decrease over time.

**Figure 2.**
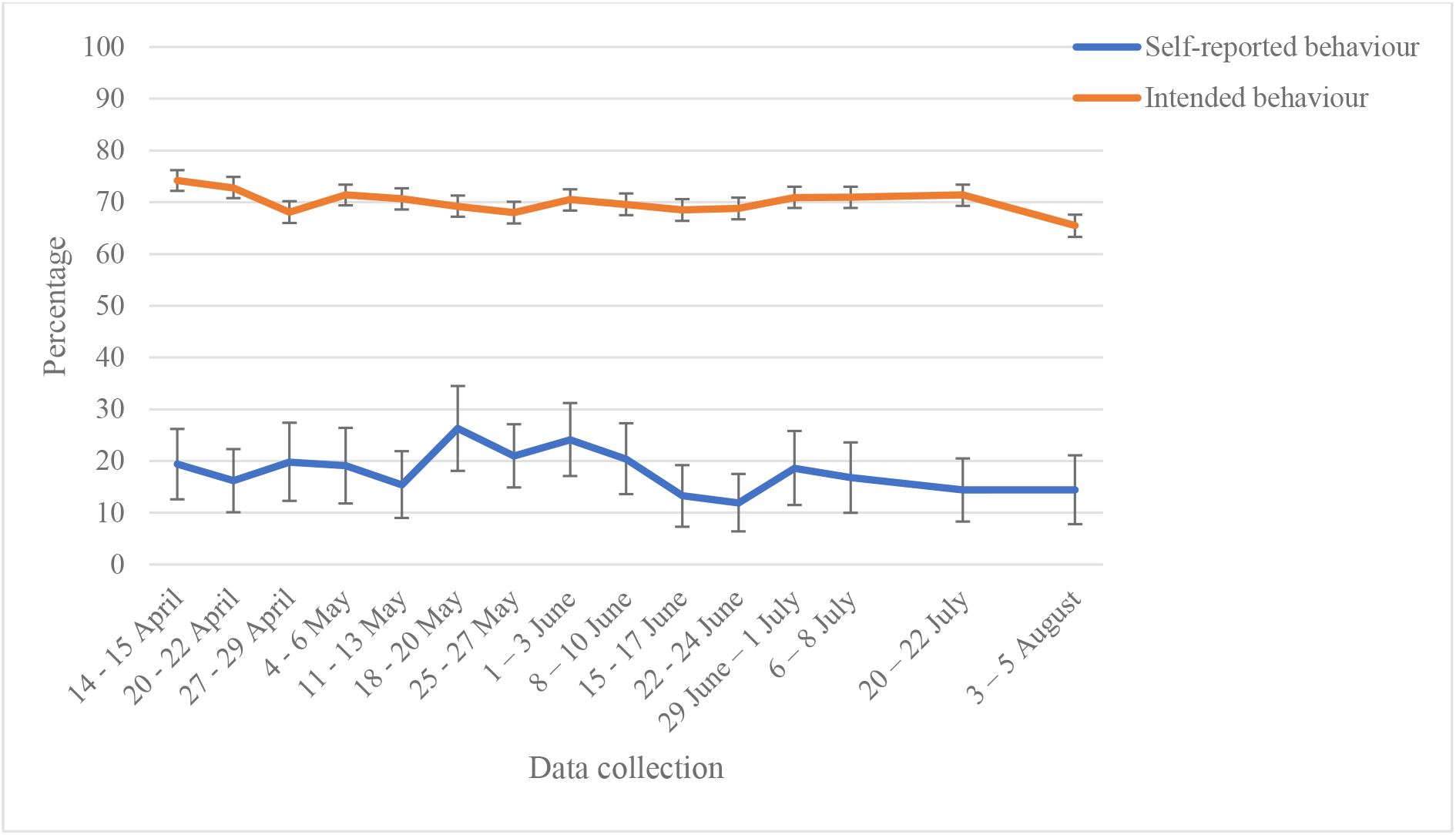
Percentage of people who reported not leaving home at all since developing COVID-19 symptoms (in those who had experienced COVID-19 symptoms in the last seven days), and who reported intending not to leave home at all if they were to develop COVID-19 symptoms (in people who had not had COVID-19 symptoms in the last seven days). Error bars are 95% confidence intervals.

The factors most strongly associated with non-adherence to self-isolation were: not knowing Government guidance about what to do if you developed COVID-19 symptoms; not identifying COVID-19 symptoms; thinking you have had COVID-19; having a dependent child in the household; and working in a key sector (see Tables 4 and 5). Self-reported reasons for not self-isolating are presented in the supplementary materials. The most common reasons were: to go to the shops for groceries/pharmacy (18.2%); because one’s symptoms got better (15.6%); and to go out for a medical need other than COVID-19 (14.9%).

### Requesting an antigen test

Of those who reported experiencing COVID-19 symptoms in the last seven days, only 11.9% (95% CI 10.1% to 13.8%) reported requesting an antigen test. While intention to request a test has increased over time, self-reported behaviour has remained relatively stable (see Figure 3).

**Figure 3.**
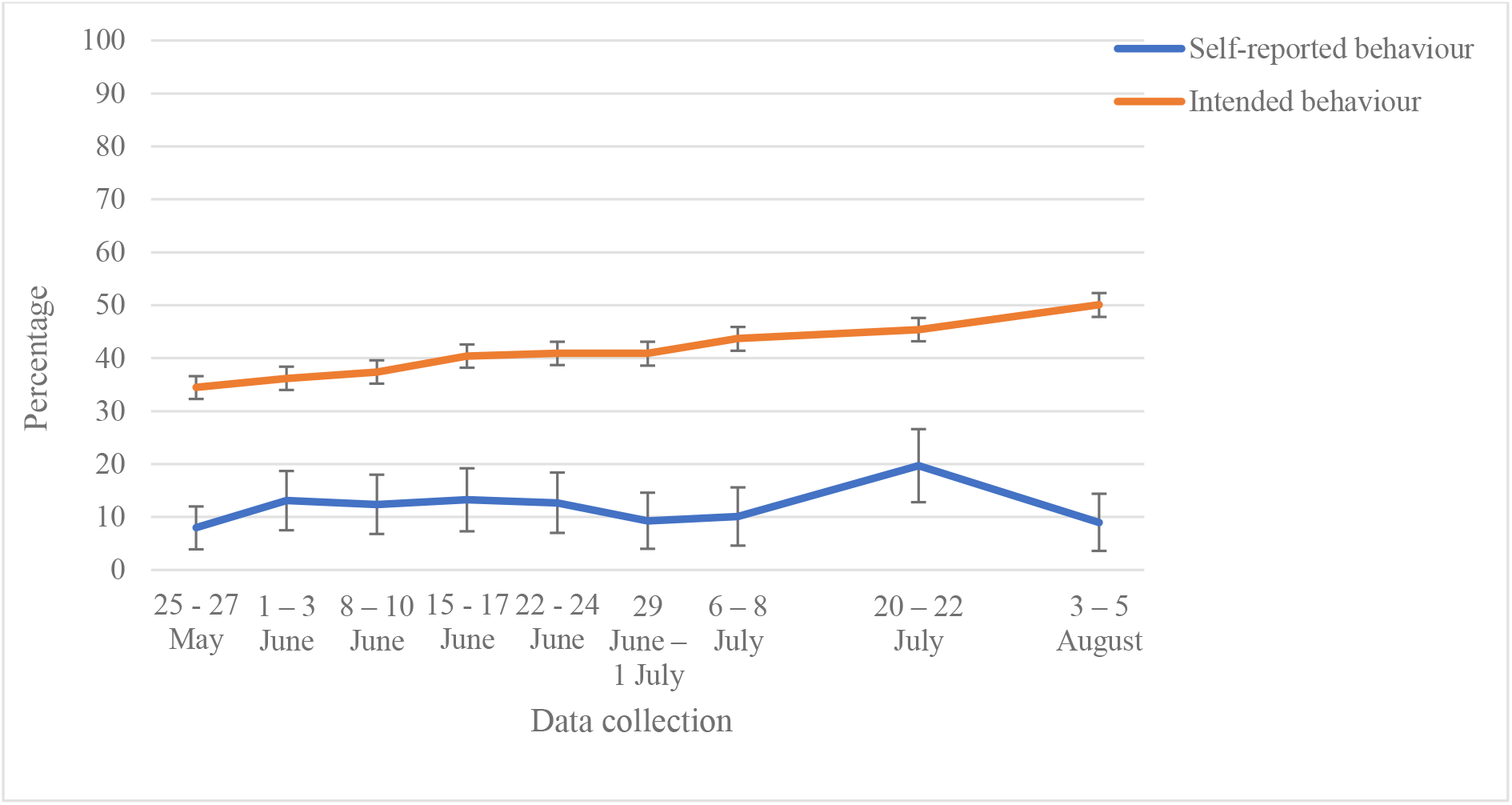
Percentage of people who reported requesting an antigen test after developing COVID-19 symptoms (in those who had experienced COVID-19 symptoms in the last seven days), and who reported intending to request an antigen test if they were to develop COVID-19 symptoms (in people who had not had COVID-19 symptoms in the last seven days). Error bars are 95% confidence intervals.

The only factor strongly associated with not requesting an antigen test was lower confidence that you could return a completed home-testing kit for COVID-19 by courier (see Tables 5 and 6). Common reasons for not requesting an antigen test included: not thinking that symptoms were due to COVID-19 (20.0%); because symptoms improved (16.1%); and because symptoms were only mild (16.0%; see supplementary materials).

### Sharing details of close contact

Of those who had not experienced COVID-19 symptoms in the last seven days, 76.1% (95% CI 75.4% to 76.8%) reported that they probably or definitely would share details of their close contacts with the NHS contact tracing service if they tested positive for coronavirus, and were prompted by the NHS contact tracing service (see Figure 4).

**Figure 4.**
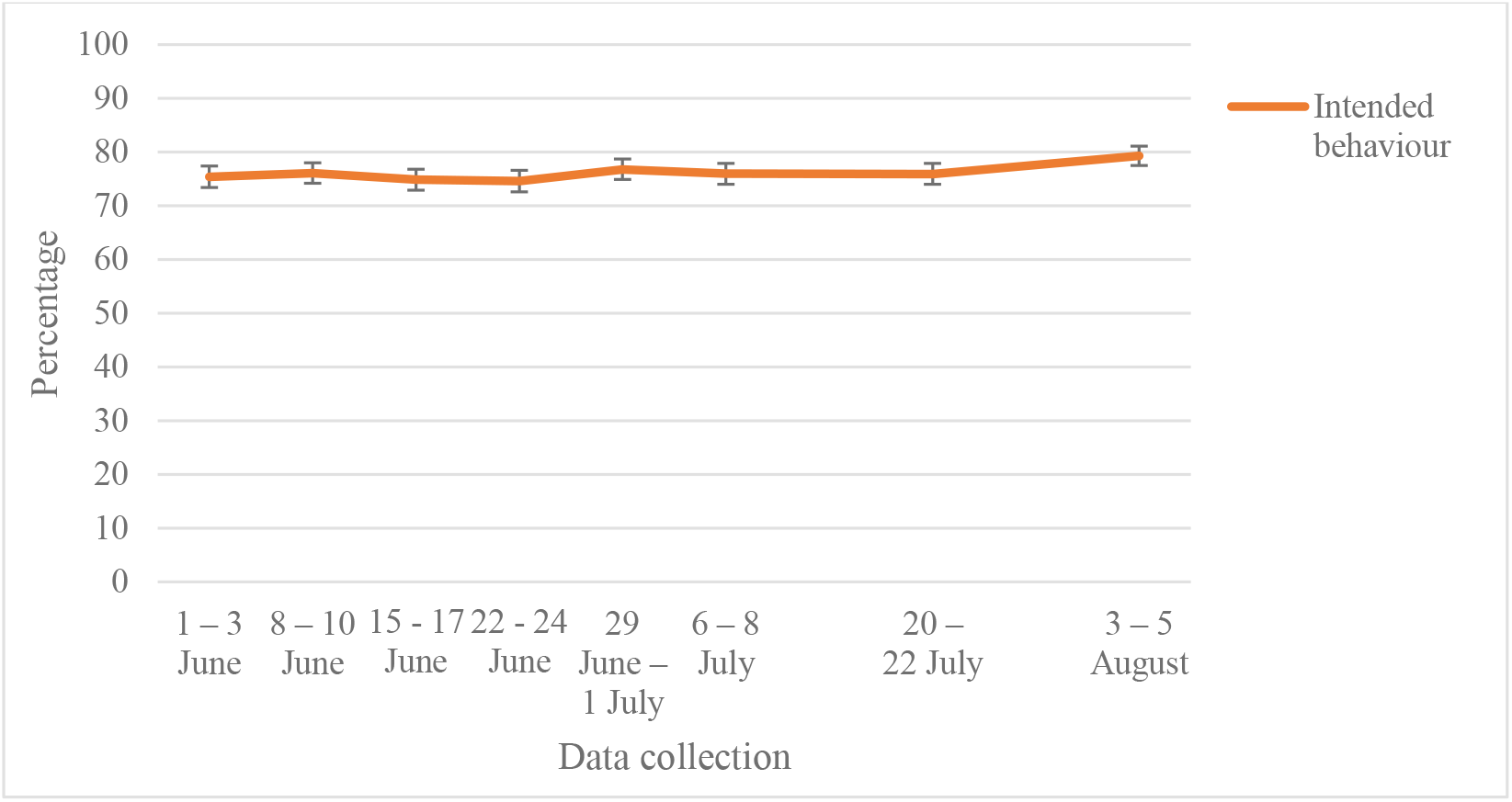
Percentage of people who reported that they probably or definitely would share details of their close contacts if contacted by the NHS contact tracing service (in people who had not had COVID-19 symptoms in the last seven days). Error bars are 95% confidence intervals.

Key factors associated with not intending to share details of close contacts if you test positive for COVID-19 were: not believing that contact tracing systems were effective at preventing the spread of COVID-19; not knowing that you can spread COVID-19 to others if you are asymptomatic; being concerned about spreading COVID-19 to someone who may be at risk; and thinking that your personal behaviour has an impact on how COVID-19 spreads (see Tables 8 and 9). The most common reasons for not intending to share details of your close contacts were: not knowing if data would be secure and confidential (18.8%); not knowing what would happen to the data (17.2%); and thinking that the contact tracing system was not accurate and reliable (14.2%; see supplementary materials).

### Quarantining after being alerted

Of those who reported having been alerted by the NHS contact tracing service and told they had been in close contact with a confirmed COVID-19 case (n=405), 10.9% (95% CI 7.8% to 13.9%) reported that they had not left home at all in the following 14 days. This figure has remained relatively stable; intention to quarantine is much higher, at around 65% (see Figure 5).

**Figure 5.**
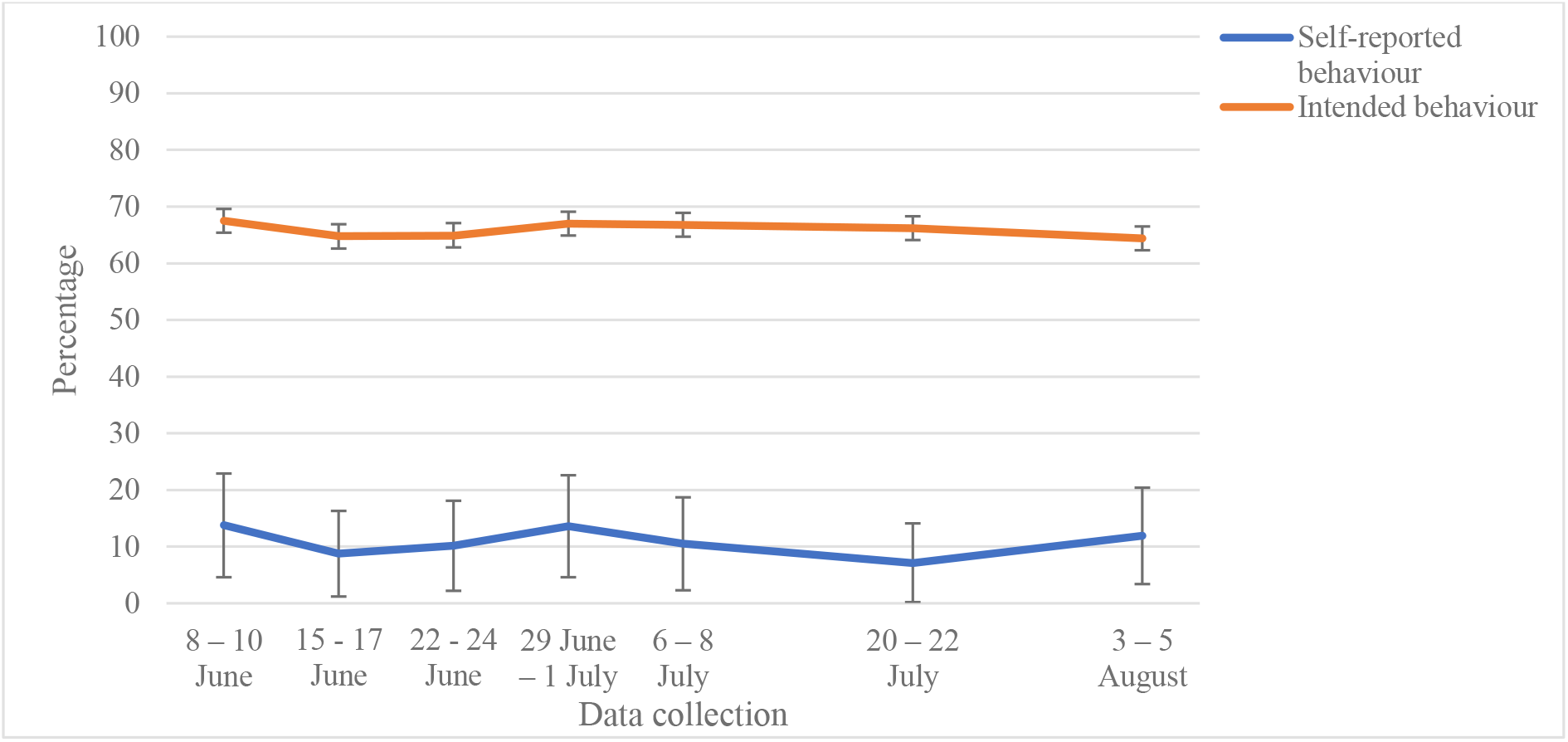
Percentage of people who reported quarantining for 14 days after being alerted that they had been in contact with a confirmed COVID-19 case by the NHS contact tracing service (the most recent time they had been alerted), and who reported intending to quarantine for 14 days if they were alerted that they had been in contact with a confirmed COVID-19 case by the NHS contact tracing service (in people who had never been alerted). Errors bars are 95% confidence intervals.

The only factor strongly associated with non-adherence to quarantine was having a dependent child in the household (see Tables 10 and 11). Key reasons given for not quarantining included: not thinking it was necessary to stay away from people outside your household as you could not stay away from people in your household (14.3%); not developing symptoms (11.9%); to go to the shops for groceries/pharmacy (10.9%); and having just finished quarantining because you had been in contact with a different confirmed COVID-19 case (10.9%; see supplementary materials).

### Factors associated with multiple outcomes

Patterns of results were similar for all outcomes. Lower adherence was associated with being male, younger age, having a dependent child in the household, lower socio-economic grade, greater hardship, and being less informed about COVID-19 and guidance to prevent the spread of the virus (e.g. not being able to identify key symptoms of COVID-19, not knowing government guidance if you were to develop symptoms of COVID-19, and disagreeing that someone can spread COVID-19 even if they are asymptomatic).

## DISCUSSION

As in other countries, the test, trace and isolate system is the cornerstone of the UK’s public health strategy for coping with the COVID-19 pandemic.(1) Its success relies on adherence to multiple behaviours at multiple stages.(6) Our data suggest that self-reported rates of adherence to isolating, testing and quarantining are currently low, as are rates of recognition of the main symptoms of COVID-19 (see Figure 6). Rates of intended behaviour are much higher than rates of self-reported behaviour. This is unsurprising.(33) The percentage of people who share details of close contacts after receiving a positive COVID-19 antigen test is therefore likely much lower than the percentage of intended sharing of contacts reported here.

Our observed rates of adherence were largely stable over time with the notable exceptions of symptom recognition, where recognition of ’new’ symptoms increased over the first one or two weeks after they were introduced, and intended (but not actual) requests for an antigen test, which have been steadily increasing. These findings are in line with other research finding low rates of symptom identification and low adherence to self-isolation in the UK.(8, 9, 16) Our estimate of the percentage of people requesting a test (11.9%) is lower than the estimate that can be derived by dividing the number of cases per day identified in the community by NHS Test and Trace (34) by the estimated daily incidence recorded by the Office of National Statistics (35) (35% for 6-13 August). This discrepancy might be accounted for by different sample biases, the probable inclusion of people in our sample with an obvious, non-COVID explanation for their symptoms, and the probable inclusion of asymptomatic cases in the NHS Test and Trace data.

Stability of the outcomes indicates that changes to messaging between March and early August have had little effect on behaviours relevant to test, trace and isolate. This suggests either that the changes introduced so far have been ineffective, the budget allocated to messaging about the test, trace and isolate system has been insufficient to allow changes to have an impact, or that the factors preventing people from engaging with behaviours are not amenable to messaging alone.

Our results suggest that financial constraints and caring responsibilities impeded adherence to self-isolation, intending to share details of close contacts, and quarantining of contacts. The disproportionate impact of the pandemic on people from lower socio-economic backgrounds and with caring responsibilities has been well-documented.(36, 37) Previous research has shown that people who have received help from others outside their household because of COVID-19 were more likely to adhere to self-isolation.(9) To encourage adherence, policies must ensure that people are adequately reimbursed for any potential losses that may arise from needing to self-isolate and facilitate practical considerations, such as shopping for groceries and medicines during self-isolation.

In terms of capability,(7) it appears that higher knowledge in general was associated with greater uptake of protective behaviours. It is impossible to disentangle causality here. People who are better informed may simply be more engaged generally in attempting to understand and tackle the pandemic, with the latter promoting adherence. Nevertheless, disseminating clear and easily understood information about the virus and how it spreads is likely to increase adherence to protective behaviours, especially where understanding is low. Motivational factors, such as perceiving measures to be effective and being confident about returning a testing kit, were associated with intending to share details of close contacts and requesting a test respectively. This is in line with research findings from the H1N1 influenza pandemic.(14) Making antigen testing as easy as possible, for example by introducing local testing sites in areas with high infection rates,(38) may increase adherence to testing. Messaging that highlights the effectiveness of adhering to each stage of the test, trace and isolate system, and that emphasises that adhering is straightforward and easy to do may further improve adherence.

With regard to demographic differences, men and younger people were less likely to adhere to steps along the test, trace and isolate pathway. Similar findings emerged during the H1N1 pandemic.(14) Gender and age differences in adherence may be caused by differences such as poorer health literacy in men and a greater desire to be active and to have contact with peer groups amongst younger age groups.(39) Working in a key sector was associated with not self-isolating. This may be because key workers have a greater financial need or feel a greater social pressure to attend work and are less likely to be able to work from home.(40) Key workers and people from minority ethnic backgrounds were less likely to identify common symptoms of COVID-19. Targeted communications to these groups may help improve adherence and increase knowledge of common symptoms of COVID-19.

We found no evidence for associations between perceived risk to oneself or to people in the UK and adherence to test, trace and isolate behaviours. We also found no evidence for an association between self-isolation and concern about spreading the virus to those at risk of complications, thinking that your personal behaviour has an impact on the spread of COVID-19, or perceived effectiveness of self-isolation. However, these factors were associated with intention to share details of close contacts. Taken together, these findings suggest either that concern about the risk of giving COVID-19 to others increases intention to adhere to protective behaviours but not actual behaviour, or that such concerns are mainly important where any inconvenience associated with the behaviour is imposed on someone else (as is the case of quarantine) rather than on you (as is the case in self-isolation).

We found an association between perceiving information from the Government to be more credible and being less likely to self-isolate and to identify COVID-19 symptoms. It may be that this association is confounded by, for example, the political orientation of participants.

Strengths of this study include large sample sizes allowing us to investigate infrequent behaviours. Inclusion of survey items in multiple waves of data collection has enabled us to track uptake of protective behaviours and knowledge over time. This study also has limitations. First, we used quota sampling to ensure that participant characteristics were representative of the UK adult population. While we cannot be sure that survey respondents are representative of the general population,(41, 42) online quota sampling is a pragmatic approach when a large, demographically representative sample must be obtained in a very short time frame during a crisis.(14, 43) Second, data were self-reported, and so could have been influenced by social desirability and recall gaps and bias. Although self-reported adherence to protective measures for COVID-19 such as social distancing is associated with real-world behaviour,(44) it is likely that rates reported here are overestimates of adherence. Third, data are cross-sectional, therefore we cannot infer causality. Fourth, although we asked participants if they had left home at all since developing COVID-19 symptoms, technically it is permissible to leave home under some circumstances, including to attend a medical appointment, get a test or if you receive a negative test result. Given that only 12% of people with symptoms reported requesting a test, we do not believe this explanation accounts for more than a small fraction of the non-adherence that we observed. Fifth, while we had a large overall sample size, numbers of participants included in analyses investigating requesting an antigen test and quarantining after being alerted were smaller, and skewed outcome responses resulted in small cell counts. We have presented these analyses for completeness, but these results should be treated with caution.

In order for the test, trace and isolate system in the UK to succeed, people must recognise the key symptoms of COVID-19, self-isolate, request a test, share details of their close contacts and quarantine if contacted. Our results indicate that approximately half of people know the symptoms of COVID-19, and that adherence to each stage of the test, trace and isolate journey is low. Policies which support people financially and practically, and providing and communicating about a testing system that is both effective and easy to access, will be key to increasing uptake. Targeted communications especially to men, younger age groups and key workers are likely to further increase uptake.

## Data Availability

No additional data are available from the authors.

## TRANSPARENCY DECLARATION

The authors affirm that the manuscript is an honest, accurate, and transparent account of the study being reported; that no important aspects of the study have been omitted; and that any discrepancies from the study as originally planned have been explained.

Surveys were commissioned and funded by DHSC, with the authors providing advice on the question design and selection. DHSC had no role in analysis, decision to publish, or preparation of the manuscript. Preliminary results were made available to DHSC and the UK’s Scientific Advisory Group for Emergencies (SAGE). DHSC requested that we delay publication until August 2020.

## DATA SHARING STATEMENT

No additional data are available from the authors.

## AUTHOR CONTRIBUTION STATEMENT

All authors conceptualised the study and contributed to survey materials. LS completed analyses with guidance from HWWP and GJR. LS and GJR wrote the first draft of the manuscript. All authors contributed to, and approved, the final manuscript. GJR is guarantor. The corresponding author attests that all listed authors meet authorship criteria and that no others meeting the criteria have been omitted.

## LICENSE

The Corresponding Author has the right to grant on behalf of all authors and does grant on behalf of all authors, a worldwide licence (<u>the BMJ license</u>) to the Publishers and its licensees in perpetuity, in all forms, formats and media (whether known now or created in the future), to i) publish, reproduce, distribute, display and store the Contribution, ii) translate the Contribution into other languages, create adaptations, reprints, include within collections and create summaries, extracts and/or, abstracts of the Contribution and convert or allow conversion into any format including without limitation audio, iii) create any other derivative work(s) based in whole or part on the on the Contribution, iv) to exploit all subsidiary rights to exploit all subsidiary rights that currently exist or as may exist in the future in the Contribution, v) the inclusion of electronic links from the Contribution to third party material where-ever it may be located; and, vi) licence any third party to do any or all of the above. All research articles will be made available on an open access basis (with authors being asked to pay an open access fee—see <u>copyright, open access, and permission to reuse</u>). The terms of such open access shall be governed by a <u>Creative Commons</u> licence—details as to which Creative Commons licence will apply to the research article are set out in our worldwide licence referred to above.

## DISSEMINATION DECLARATION

Dissemination of survey results to participants is not possible due to the anonymous nature of data collection.

